# Genetic modifiers and ascertainment drive variable expressivity of complex disorders

**DOI:** 10.1101/2024.08.27.24312158

**Authors:** Matthew Jensen, Corrine Smolen, Anastasia Tyryshkina, Lucilla Pizzo, Deepro Banerjee, Matthew Oetjens, Hermela Shimelis, Cora M. Taylor, Vijay Kumar Pounraja, Hyebin Song, Laura Rohan, Emily Huber, Laila El Khattabi, Ingrid van de Laar, Rafik Tadros, Connie Bezzina, Marjon van Slegtenhorst, Janneke Kammeraad, Paolo Prontera, Jean-Hubert Caberg, Harry Fraser, Siddhartha Banka, Anke Van Dijck, Charles Schwartz, Els Voorhoeve, Patrick Callier, Anne-Laure Mosca-Boidron, Nathalie Marle, Mathilde Lefebvre, Kate Pope, Penny Snell, Amber Boys, Paul J. Lockhart, Myla Ashfaq, Elizabeth McCready, Margaret Nowacyzk, Lucia Castiglia, Ornella Galesi, Emanuela Avola, Teresa Mattina, Marco Fichera, Maria Grazia Bruccheri, Giuseppa Maria Luana Mandarà, Francesca Mari, Flavia Privitera, Ilaria Longo, Aurora Curró, Alessandra Renieri, Boris Keren, Perrine Charles, Silvestre Cuinat, Mathilde Nizon, Olivier Pichon, Claire Bénéteau, Radka Stoeva, Dominique Martin-Coignard, Sophia Blesson, Cedric Le Caignec, Sandra Mercier, Marie Vincent, Christa Martin, Katrin Mannik, Alexandre Reymond, Laurence Faivre, Erik Sistermans, R. Frank Kooy, David J. Amor, Corrado Romano, Joris Andrieux, Santhosh Girirajan

## Abstract

Variable expressivity of disease-associated variants implies a role for secondary variants that modify clinical features. We assessed the effects of modifier variants towards clinical outcomes of 2,252 individuals with primary variants. Among 132 families with the 16p12.1 deletion, distinct rare and common variant classes conferred risk for specific developmental features, including short tandem repeats for neurological defects and SNVs for microcephaly, while additional disease-associated variants conferred multiple genetic diagnoses. Within disease and population cohorts of 773 individuals with the 16p12.1 deletion, we found opposing effects of secondary variants towards clinical features across ascertainments. Additional analysis of 1,479 probands with other primary variants, such as 16p11.2 deletion and *CHD8* variants, and 1,084 without primary variants, showed that phenotypic associations differed by primary variant context and were influenced by synergistic interactions between primary and secondary variants. Our study provides a paradigm to dissect the genomic architecture of complex disorders towards personalized treatment.

## INTRODUCTION

As large-scale sequencing studies uncover increasingly complex links between genomic variants and heritable disorders, identifying the genetic etiology in an affected individual has become more challenging^1^. In contrast to Mendelian disorders caused by single genes, many disorders are characterized by extensive phenotypic heterogeneity, where individuals with the same variant exhibit a range of phenotypes with variable penetrance and expressivity^2,3^. Some instances of phenotypic heterogeneity can be explained by multiple genetic diagnoses, where more than one pathogenic variant contributes to largely independent disorders in the same individual^4^. These variants can even synergistically contribute to new phenotypes not associated with the individual variants, such as seizures observed among individuals with variants in both *MKS1* (associated with Meckel-Gruber syndrome) and *BBS1* (associated with Bardet-Biedl syndrome)^5^. Other cases of phenotypic heterogeneity could occur when the clinical features of causal variants are modified by secondary variants that do not cause disease themselves^6^. For example, rare variants in histone modifier genes were enriched among individuals with the 22q11.2 deletion who exhibited variably expressive congenital heart defects^7^. The complexity increases exponentially for neurodevelopmental disorders, where the combined effects of primary and secondary variants with differing frequency and effect sizes explain their broad heterogeneity^8,9^. For example, recent studies have found significant contributions of polygenic risk from common variants towards phenotypes of individuals with pathogenic copy-number variants (CNVs)^10,11^, such as schizophrenia risk in individuals with 22q11.2 deletion^12–17^. Further, variable expressivity could also be explained by cohort ascertainment, as many pathogenic variants are enriched among individuals across disease ascertainments and lead to medical consequences in the general population or healthy-biased cohorts^18–22^. For example, the autism-associated 16p11.2 deletion^23^ is also associated with obesity, musculoskeletal, pulmonary, hematologic, and renal features in the general population^24,25^. This complex interplay necessitates a systematic assessment to fully understand which modifier variants contribute to specific phenotypes when ascertained for the same primary variant.

Rare recurrent CNVs represent excellent models to study variable expressivity, as the large number of duplicated or deleted genes increases the likelihood of interactions with modifiers elsewhere in the genome^3,26^. For example, the rare heterozygous 520-kbp 16p12.1 deletion (hg18/NCBI36 when originally reported; currently maps to 16p12.2 based on hg19/GRCh37) is enriched among children with severe neurodevelopmental features and is inherited in >90% of cases from a parent who manifests milder psychiatric and cognitive features^27–29^. The phenotypic manifestation among individuals with this deletion differs based on cohort ascertainment. For instance, the deletion was originally described in children with developmental delay^27^, but studies from the general population also identified associations with multiple psychiatric and cognitive features^22,30–32^. Thus, the 16p12.1 deletion is an ideal paradigm for studying the effects of modifier variants on the clinical trajectory of a primary variant. We previously found that severely affected children with the deletion have a global increase in rare variant burden compared to parents with the deletion, and these trends are consistent for other primary variants^27,28,33^. Our findings suggested a “multi-hit” model for complex disease etiology, where a primary variant sensitizes an individual for disease and the clinical outcome is determined by other “hits” elsewhere in the genome^3^. However, it is not completely understood how specific variant classes of differing effect size and frequency modify clinical features across different ascertainment and primary variant contexts.

Here, we performed deep clinical and quantitative phenotyping and comprehensive analysis of genomic data for 2,252 individuals with primary variants from diverse disease and population-based cohorts (**Fig. 1**). We systematically dissected the roles of multiple secondary variant classes towards developmental features in 132 families with the 16p12.1 deletion and expanded our analysis to uncover phenotypic associations in 773 16p12.1 deletion individuals from disease cohorts and healthy populations as well as 1,479 autism probands who carry a range of other primary variants and 1,084 autism probands without primary variants. Our results show that variant-phenotype associations are dependent on both the primary and secondary variant context as well as cohort ascertainment (**Fig. 1**), allowing for more accurate dissection of the genetic etiology of variably expressive traits associated with pathogenic variants.

**Figure 1.**
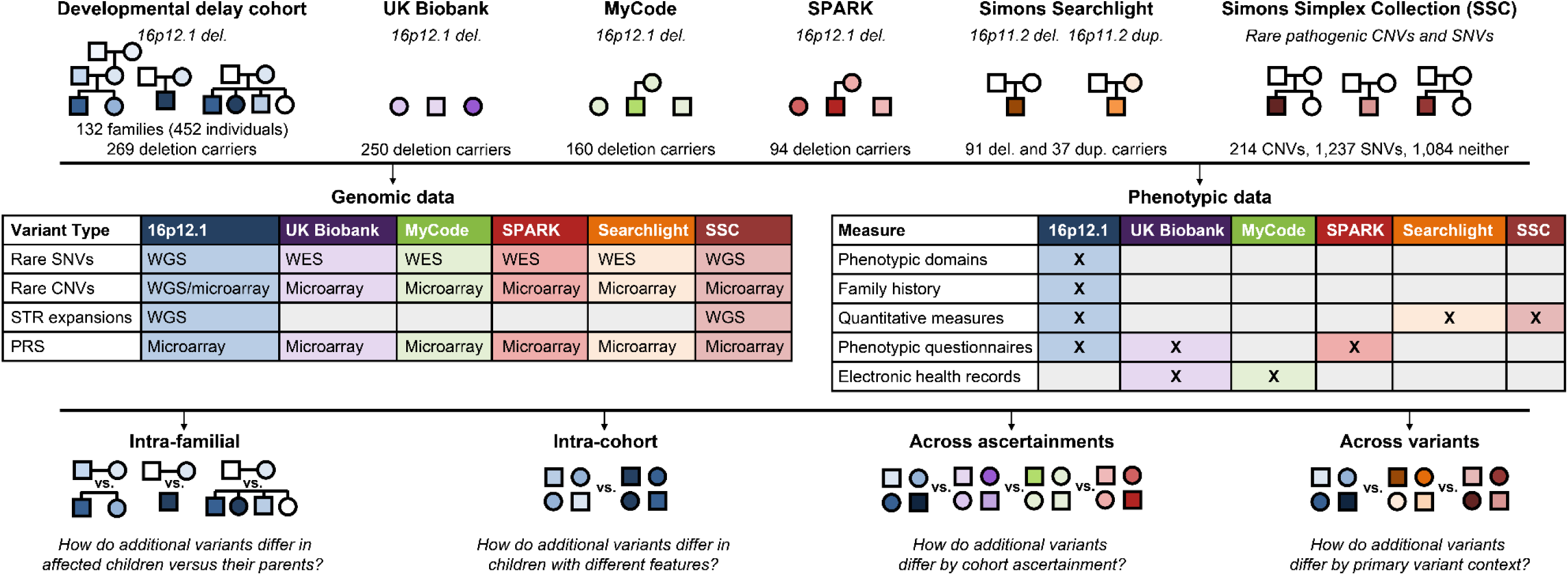
Overview of variant-phenotype analyses in 2,252 individuals with primary pathogenic variants. We assessed associations between variant classes and clinical phenotypes in six cohorts of individuals and families with primary variants. We directly recruited and assessed 132 families with the 16p12.1 deletion primarily ascertained for children with developmental delay (DD) (also including ten individuals from eight families from the Estonian Biobank not ascertained for DD). We further assessed 16p12.1 deletion carriers from cohorts with different ascertainments, including healthy volunteer-biased (UK Biobank), clinically-derived (MyCode), and single-disorder (SPARK, for autism) ascertainments. We finally assessed probands ascertained for autism with various primary pathogenic variants, including the 16p11.2 deletion or duplication (Simons Searchlight) and other large CNVs or rare SNVs in neurodevelopmental genes (SSC). We note that 100 probands in SSC have both pathogenic SNVs and CNVs and are included in both categories. Within and across these cohorts, we identified associations between up to 17 classes of rare and common variants (identified from WGS, WES, and microarrays) with phenotypic features from deep clinical datasets and electronic health records.

## RESULTS

### Variability of clinical features in 16p12.1 deletion

We recruited a cohort of 442 individuals from 124 families with the 16p12.1 deletion (“DD cohort”), including multi-generational families, primarily ascertained for having children with developmental delay (DD) (**Fig. 1**). We analyzed phenotypes from medical records, family interviews, and online assessments for quantitative traits, such as non-verbal IQ^34^ and social responsiveness scores for autism-related social traits (SRS^35^) (**Table S1A**). In total, 93% of probands (84/90) inherited the deletion from a parent, with a slight bias towards maternal inheritance (48/84, 57%), and 70% (87/124) of probands were male. Probands with the deletion exhibited clinical features grouped across six broadly defined phenotypic domains, including intellectual disability/developmental delay (ID/DD), behavioral, psychiatric, nervous system, congenital, and growth/skeletal abnormalities (**Fig. 2A, Table S1A-B**). Probands also showed a higher number of childhood developmental and behavioral features (i.e., increased complexity scores, see *Methods*) compared to their siblings and cousins, while carrier siblings and cousins manifested more features than non-carriers (**Fig. 2A**). Parents who transmitted the deletion (“carrier parents”) often manifested milder cognitive or psychiatric phenotypes (**Fig. 2B)**.

**Figure 2.**
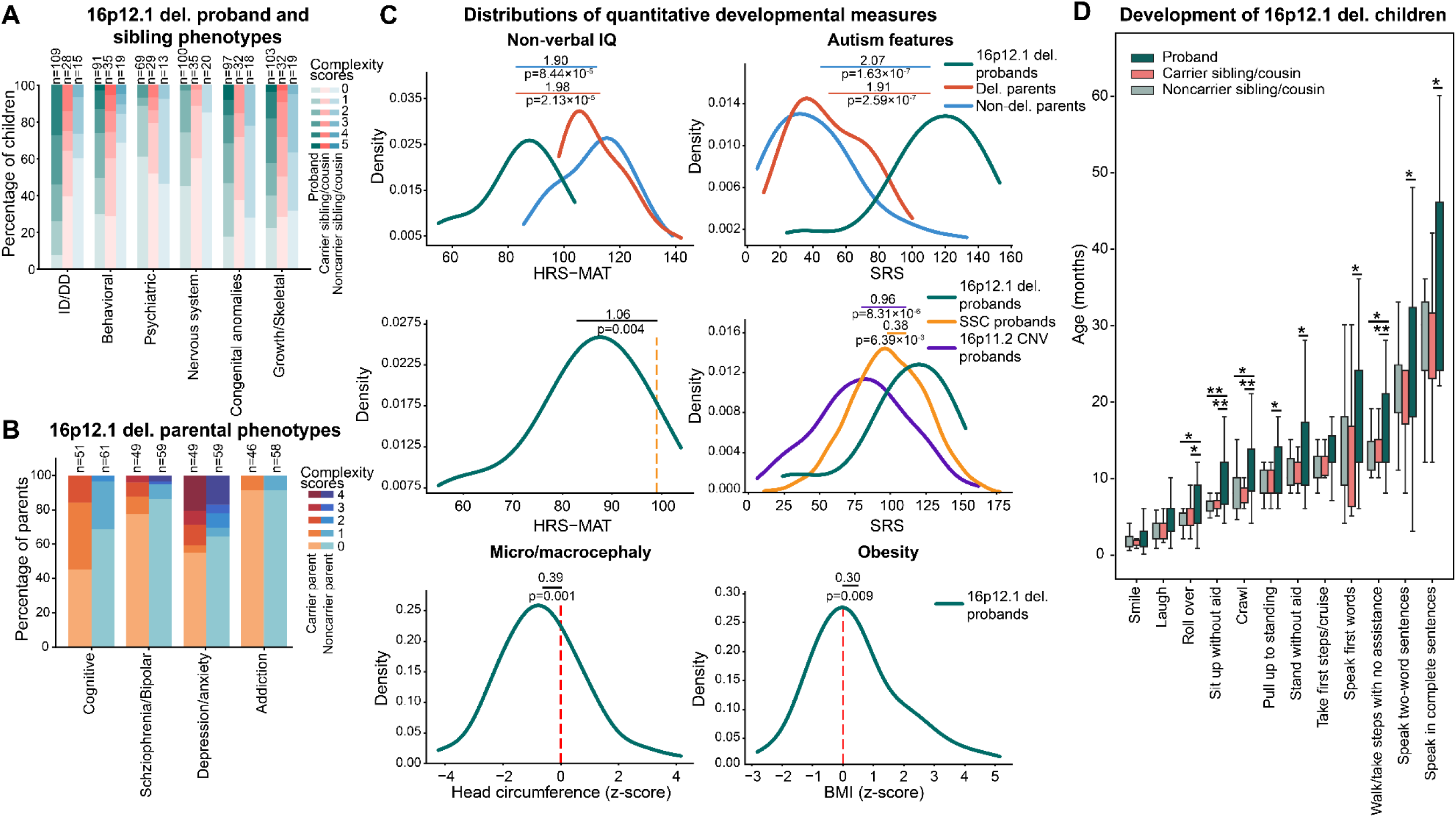
Variably expressive phenotypes of family members with the 16p12.1 deletion. (**A**) Distribution of complexity scores for six phenotypic domains in probands (n=69-109), carrier siblings and cousins (n=28-35), and noncarrier siblings and cousins (n=13-20) in 16p12.1 deletion families (numbers vary due to clinical data availability). Complexity scores were determined by identifying the number of clinical features manifested within each phenotypic domain (see Methods). **(B)** Distribution of complexity scores for four phenotypic domains in carrier parents (orange, n=46-51, orange) and non-carrier parents (blue, n=58-61) of 16p12.1 deletion probands. **(C)** Distributions of quantitative phenotypes observed in 16p12.1 deletion probands. Top plots show the distribution of non-verbal IQ (HRS-MAT) and social responsiveness scores (SRS) in probands (green, n=10-27) compared to carrier (red, n=17-21) and non-carrier parents (blue, n=20-26). Middle plots compare the same scores in probands to the score for probands in the SSC cohort (SRS n=2,844, yellow; HRS-MAT mean derived from ^34^) and probands with the 16p11.2 deletion or duplication from Simons Searchlight (n=139, purple). Bottom plots show the distribution of head circumference (n=64) and BMI z-scores (n=67) in deletion probands; red vertical lines represent the general population mean (i.e. z-score=0). P-values from Mann Whitney tests or one-sample t-tests. Individual scores for 16p12.1 deletion probands and parents are listed in **Table S1A**. **(D)** Distribution of the age of attainment for developmental milestones in probands (n=13-33), carrier siblings and cousins (n=16-18), and noncarrier siblings and cousins (n=11-15). One-tailed t-test, *p≤0.05, **Benjamini-Hochberg FDR≤0.05.

Probands had a 1.98 SD decrease in non-verbal IQ (p=2.13×10^−5^) and a 1.91 SD increase in SRS (p=2.59×10^−7^) compared with their carrier parents (**Fig. 2C, Table S2A**). The average IQ score among 16p12.1 deletion probands was 1.06 SD lower than all probands ascertained for autism from the Simons Simplex Collection^34^ (SSC) (p=0.004). The average SRS of 16p12.1 deletion probands was 0.96 SD higher than probands with 16p11.2 deletions or duplications from the Simons Searchlight cohort (p=8.31×10^−6^) and 0.38 SD higher than SSC probands (p=6.39×10^−3^) (**Fig. 2C, Table S2A**). Beyond psychiatric traits, 16p12.1 deletion probands also showed decreased head size (p=0.001) and increased body mass index (BMI, p=0.009) (**Fig. 2C, Table S2A**). Finally, consistent with their ascertainment, probands exhibited significant delays in several developmental milestones^36^ compared to their siblings (p<0.05) (**Fig. 2D, Table S2B**). Thus, our cohort represents families ascertained for probands who exhibit a range of developmental features, including more severe IQ and social responsiveness deficits than probands ascertained for autism or the 16p11.2 deletion.

### Patterns of secondary variants within and across families

Using WGS and microarray data, we evaluated 17 classes of secondary variants, including rare coding SNVs (missense and splice variants with CADD Phred scores ≥ 25 and LOF variants), non-coding SNVs (5’ untranslated region [UTR], promoter [1kb upstream of transcription start site], and enhancer [variants in regions with enhancer chromatin signatures in fetal brain] variants), rare CNVs (deletions and duplications), and short tandem repeat expansions (STRs, defined as repeat length ≥2SD than the cohort mean), a subset of which disrupted genes under evolutionary constraint^37^ (defined as LOEUF<0.35 and referred to as “(LF)” variants) (**Table S1A**). We also calculated polygenic risk scores (PRS) for four psychiatric features, including schizophrenia, intelligence, educational attainment, and autism^38–41^. These secondary variants could contribute to independent diagnoses from the 16p12.1 deletion, additively contribute to the same phenotypes as the deletion, or synergistically modify the phenotypes of the deletion. We first assessed whether probands carried additional pathogenic CNVs^33^ or secondary variants that were also present in ClinVar^42^ or in genes present in clinically relevant databases, such as Online Mendelian Inheritance in Man^43^ (OMIM), Developmental Brain Disorder database^44^ (DBD), and SFARI Gene^45^. Overall, 58% of probands (57/99) had at least one such variant, including 19% (19/99) of probands who had ClinVar-defined pathogenic variants (**Fig. S1A, Table S1D**). A subset of these cases represented probands with multiple genetic diagnoses^4^. For instance, one proband had a loss-of-function (LOF) variant in *KMT2A* and manifested Wiedemann-Steiner syndrome features, including ID/DD, dysmorphic features, and hypertrichosis^46^. Another proband with an LOF variant in *DMD* showed expected hypotonia and muscular abnormalities as well as ID/DD and craniofacial defects^28^. Additionally, we found 17 probands with STR expansions in spinocerebellar ataxia genes^47^ such as *ATXN7* and *CACNA1A*^45^. Although these probands had fewer repeats than the clinical threshold for ataxia, 64.7% (11/17) of them manifested nervous system phenotypes. We also identified a deleterious missense variant in *POLR3E* on the non-deleted allele in a proband with global developmental delay and multiple congenital defects (such as bilateral club feet and natal teeth).

We next sought to identify patterns of secondary variants in probands compared to their parents (**Fig. S1B**). Probands carried more coding (LF) SNVs (union of missense, LOF, and splice variants) (p=0.041) and missense (LF) variants (p=0.017), as well as increases in non-coding SNVs in 5’ UTRs (p=0.045), compared to their carrier parents (**Fig. 3A, Fig. S1C, Table S2C**). Probands also carried higher schizophrenia polygenic risk than their carrier parents (p=0.009), showing that polygenic risk may also contribute to the features observed among 16p12.1 deletion probands (**Fig. 3A, Fig. S1C, Table S2C**). Except for an increase in LOF (LF) variants (p=0.039), no differences across other variant classes were observed in probands compared to non-carrier parents (**Fig. 3A, Table S2C**). This is consistent with a model in which the deletion and secondary variants are transmitted in specific patterns that lead to different clinical outcomes in probands. In fact, assessment of multi-generational families showed that variant burden tends to compound over generations towards increased phenotypic severity. For example, in one multi-generational family, the carrier grandparent had mild cognitive features, while the carrier parent manifested psychiatric features and the proband had neurodevelopmental features (**Fig. 3B**). This increase in phenotypic severity corresponded with an increase in the burden of multiple rare variant classes across generations, akin to the genetic anticipation observed for certain Mendelian disorders.

**Figure 3.**
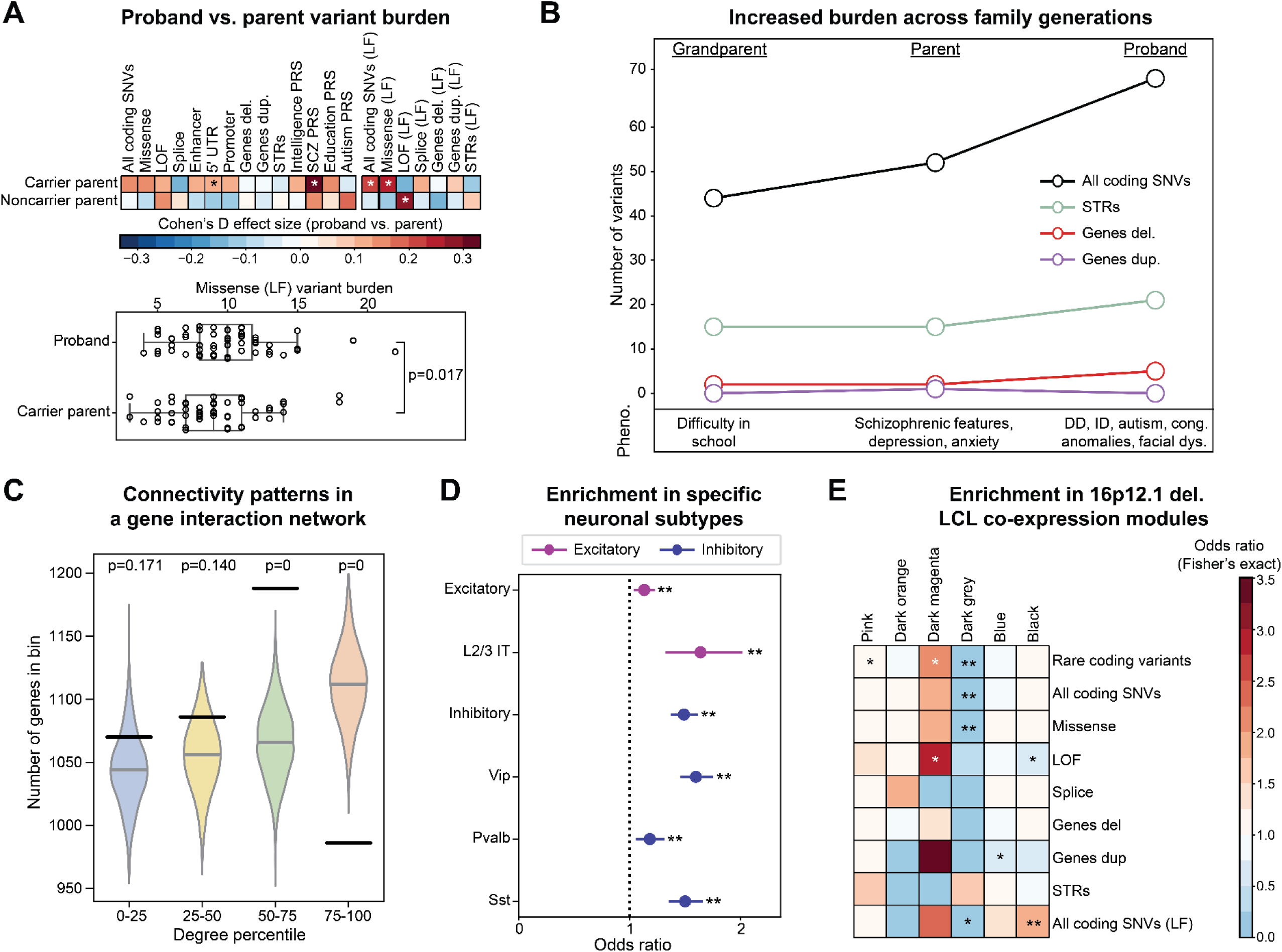
Secondary variants contribute to phenotypic variability within 16p12.1 deletion families. (**A**) Cohen’s D effect sizes (top) for changes in secondary variant burden (i.e. rare variant burden or PRS) between probands and their carrier or noncarrier parents (n=49-54 pairs). *p≤0.05, paired one-tailed (rare variant classes) or two-tailed (PRS) t-test. Red indicates increased burden in probands relative to their parents. Boxplot (bottom) highlights increased burden of missense (LF) variants between probands and carrier parents. **(B)** Increased burden of rare variants corresponds with more severe clinical features across successive generations of 16p12.1 deletion carriers in a multi-generational family. **(C)** Distribution of genes by average connectivity (degree) within a brain-specific interaction network, binned into quartiles from 1000 simulations of randomly selected gene sets. Black lines represent the observed number of genes with secondary variants in 16p12.1 deletion probands in each degree quartile. Empirical p-values derived from simulation distributions. **(D)** Enrichment of genes with secondary SNVs in 16p12.1 deletion probands for genes preferentially expressed in neuronal classes (excitatory and inhibitory) and sub-classes (colored by main class) in the adult motor cortex. Fisher’s exact test, **Benjamini-Hochberg FDR≤0.05. Full results are listed in **Table S2E**. **(E)** Enrichment of genes with secondary variants in probands for six gene co-expression modules identified from WGCNA analysis of lymphoblastoid cell lines (LCL) from individuals with the 16p12.1 deletion^54^. Fisher’s exact test, *p≤0.05, **Benjamini-Hochberg FDR≤0.05.

We further profiled the putative functions of secondary variants and found that missense variants were enriched for brain-expressed, constrained, and post-synaptic density genes (FDR≤7.41×10^−9^)^37,48^, while genes with LOF variants were depleted for these gene sets (FDR≤0.007) (**Fig. S2A, Table S2G**). This suggests that LOF variants in essential genes may not be tolerated, particularly in the presence of the deletion, while less severe variants in these genes may contribute to neurodevelopmental disorders seen in probands^26^. As further evidence of this, secondary variants were enriched (empirical p=0.000) for genes with intermediate connectivity within a brain-specific gene interaction network^49,50^ but depleted for genes with high network connectivity, which typically represent essential genes across species^51^ (empirical p=0.000; **Fig. 3C, Fig. S2B, Table S2D**). Secondary variant genes were also preferentially expressed in several brain regions during early and mid-fetal development^52^, including the frontal cortex (FDR≤0.05) and hippocampus (FDR=1.73×10^−5^) (**Fig. S2C, Table S2H**). SNVs in particular were enriched for genes preferentially expressed across multiple neuronal classes in the adult motor cortex^53^, including excitatory (FDR=0.013) and inhibitory (FDR=3.82×10^−23^) neurons (**Fig. 3D, Table S2E**). All coding SNVs (LF) were also enriched for genes co-expressed with 16p12.1 deletion genes (black module, FDR=0.016) in lymphoblastoid cell lines (LCL) derived from a subset of 19 individuals with the deletion^54^ (**Fig. 3E, Table S2F**). Overall, our results indicate that a diverse range of biologically relevant modifiers contribute to variable phenotypes in probands with 16p12.1 deletion.

### Distinct secondary variant classes contribute to specific phenotypic outcomes

We next used a series of logistic regression models to assess contributions of rare variant classes and PRS towards individual phenotypic domains. Overall, rare coding variants contributed to nervous system (logOR=0.640, p=0.032) and growth/skeletal features (logOR=0.941, p=0.004) (**Fig. 4A, Table S3A**). Specifically, STRs were associated with nervous system features (logOR=0.596, p=0.036) while SNVs were associated with growth/skeletal features (logOR=0.962, p=0.004) (**Fig. 4A, Fig. S3A, Table S3A**). In contrast, schizophrenia PRS was negatively associated with behavioral phenotypes (logOR=-0.563, p=0.046) (**Fig. 4A, Table S3A**). Combined variant models explained an average of 8% variance (McFadden’s pseudo-R^2^; range 2% to 14%) for each phenotypic domain, and showed better performance than models built using individual variant classes (average of 2% explained variance) (**Fig. S3B, Table S3A**). These estimates further highlighted the specificity of variant-phenotype associations; for example, STRs (LF) explained 12% of variance in nervous system defects but less than 4% of variance for other features (**Fig. S3B, Table S3A**). Orthogonal burden tests also identified fewer rare variants in enhancers, promoters, and 5’ UTR elements (p≤0.012) as well as increased autism PRS (p=0.028) among probands with psychiatric features (**Fig. S3C, Table S3D**). These results suggest that the modifying roles of different secondary variant classes vary across specific phenotypes, with PRS in particular modulating behavioral features.

**Figure 4.**
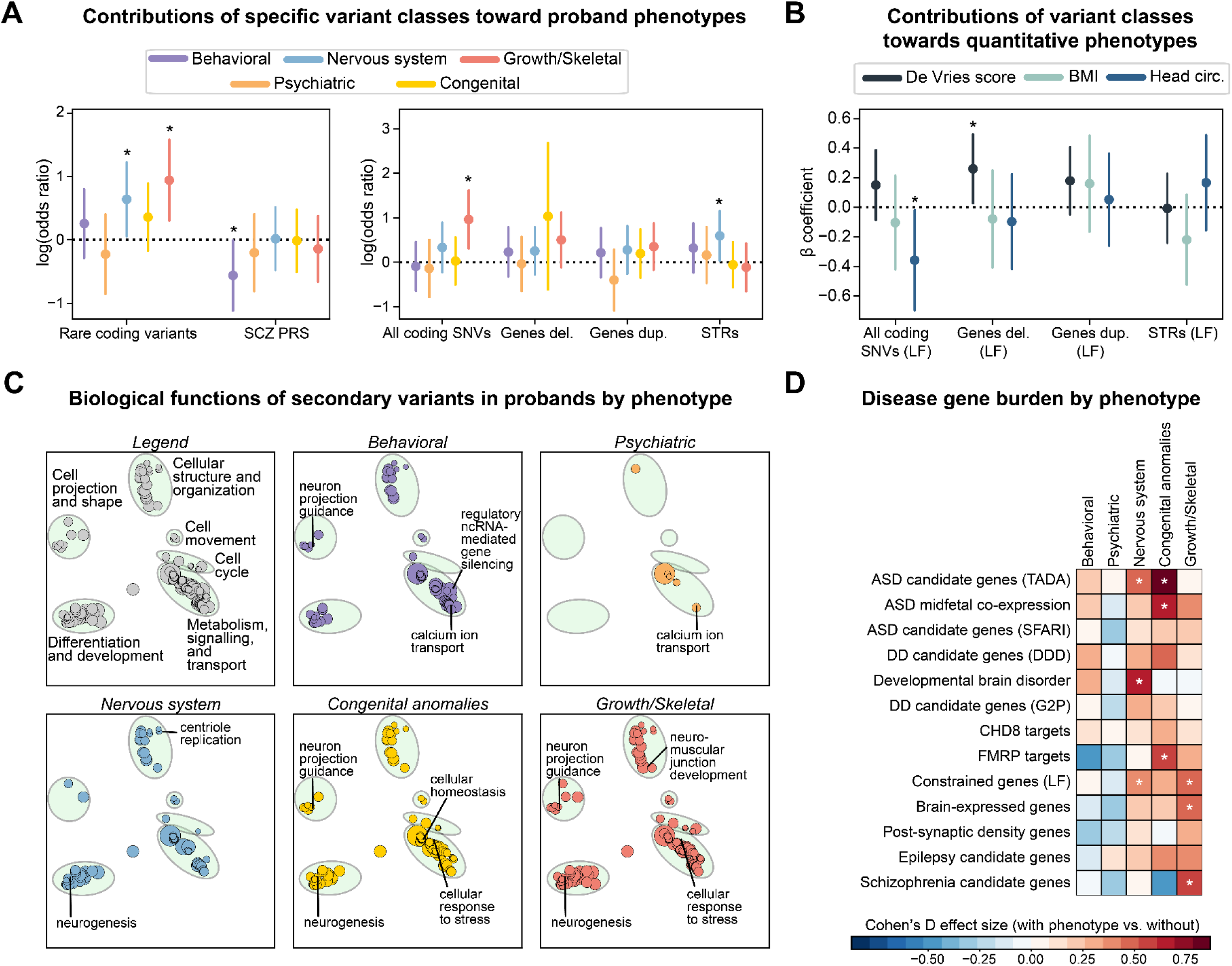
Secondary variant associations for phenotype domains of the 16p12.1 deletion. (**A**) Forest plots show log-scaled odds ratios from logistic regression models for secondary variant burden in 16p12.1 deletion probands with higher complexity scores for five phenotypic domains, compared with probands with lower complexity scores (n=47-71). *p≤0.05. Model results for variants (LF) are shown in **Fig. S3A**. **(B)** Forest plots show β coefficients from linear regression models for secondary variant burden in genes under evolutionary constrain (LF genes) towards quantitative phenotypes in deletion probands (n=43-76). *p≤0.05. Model results for variants without LF filter are shown in **Fig. S3A**. **(C)** Gene Ontology (GO) biological process terms enriched among secondary variants in probands with each phenotypic domain. Circles represent individual GO terms, clustered based on semantic similarity into broad categories (green ovals, as defined in the “legend” plot). Size of each circle represents the number of genes in each term, such that broader terms are larger. Colored circles in each plot indicate significant enrichment of the GO terms for the given phenotype. **(D)** Changes in burden of secondary variants disrupting sets of genes involved with neurodevelopmental disease and related functions (see Methods) in probands with phenotypic domains (n=23-67) compared to probands without each domain (n=12-36). *p≤0.05, one-tailed t-test.

Linear regression models testing specific variant classes towards quantitative traits revealed negative associations of head circumference z-scores with SNVs (LF) (β=-0.357, p=0.039) (**Fig. 4B, Fig. S3A, Table S3A**). Secondary CNVs were associated with increased de Vries scores, a quantitative assessment for global developmental features^55^ (deletions: β=0.288, p=0.013; duplications: β=0.246, p=0.030) (**Fig. 4B, Fig. S3A, Table S3A**). Correlation analyses revealed that intelligence and educational attainment PRS were positively correlated with head circumference (education r=0.318, p=0.026; intelligence r=0.287, p=0.045), while duplications (LF) were negatively correlated with social responsiveness deficits (r=-0.605, FDR=0.030) (**Fig. S3D**, **Table S3B**).

Secondary variants in probands ascertained for the same phenotypes showed specific enrichments for biological function, including neuromuscular junction development genes in probands with growth/skeletal defects and axonogenesis-related genes in probands with behavioral and nervous system features (**Fig. 4C, Table S3B**). Additionally, probands with specific phenotypes showed increased burden of rare variants in key neuronal genes, such as *FMRP*-binding targets^48^ (p=0.050) in probands with congenital anomalies and candidate autism^45^ (p=0.014) and developmental brain disorder genes^44^ (p=0.001) in probands with nervous system defects (**Fig. 4D, Table S3C**). Overall, our findings indicate that the disruption of distinct biological functions and molecular pathways by secondary variants may underlie specific phenotypic features of individuals with the deletion.

### Differing ascertainments confer distinct genotype-phenotype patterns

Clinical outcomes associated with the same genetic variant may vary across cohorts with different ascertainments, especially for cohorts composed of affected individuals compared to those drawn from the general population^56,57^. We sought to compare the phenotypic effects of the 16p12.1 deletion in 757 individuals with complete phenotypic data, including 253 pediatric and adult carriers from the DD cohort and three independent cohorts with distinct ascertainments: SPARK (n=94), where families were ascertained for probands with autism features^58^, and two population-based cohorts^59,60^, the healthy-biased UK Biobank (UKB; n=250) and the hospital-derived Geisinger MyCode (MyCode; n=160) (**Fig. 1**). Assessment of UKB individuals with the deletion showed enrichment for a variety of clinical phenotypes within electronic health record (EHR) data, including circulatory, endocrine, and genitourinary features (n=3,488, FDR≤0.004) (**Fig. S4A, Table S5H**). PheWAS analysis further revealed enrichment of obesity– and kidney-related features, including type 2 diabetes and hypertension (n=99,363-255,262, p≤3.48×10^−7^) (**Fig. S4B, Table S5I**). This pattern of obesity-related features is in line with the pattern of increased BMI we observed in probands in the DD cohort (**Fig. 2C**). To more directly compare phenotype prevalence across cohorts, we next harmonized EHR and clinical questionnaire responses (**Table S4A**). As expected, the prevalence of neuropsychiatric phenotypes in both pediatric and adult deletion carriers varied across the cohorts (**Fig. 5A**, **Fig. S4C-D**, **Table S4B**). For example, we found increased anxiety symptoms in adults from the DD cohort compared to UKB (p=4.18×10^−4^) (**Fig. 5A**, **Table S4B**), likely reflecting biases due to ascertainment for severely affected family members in the DD cohort compared to healthy volunteers in UKB^61^.

**Figure 5.**
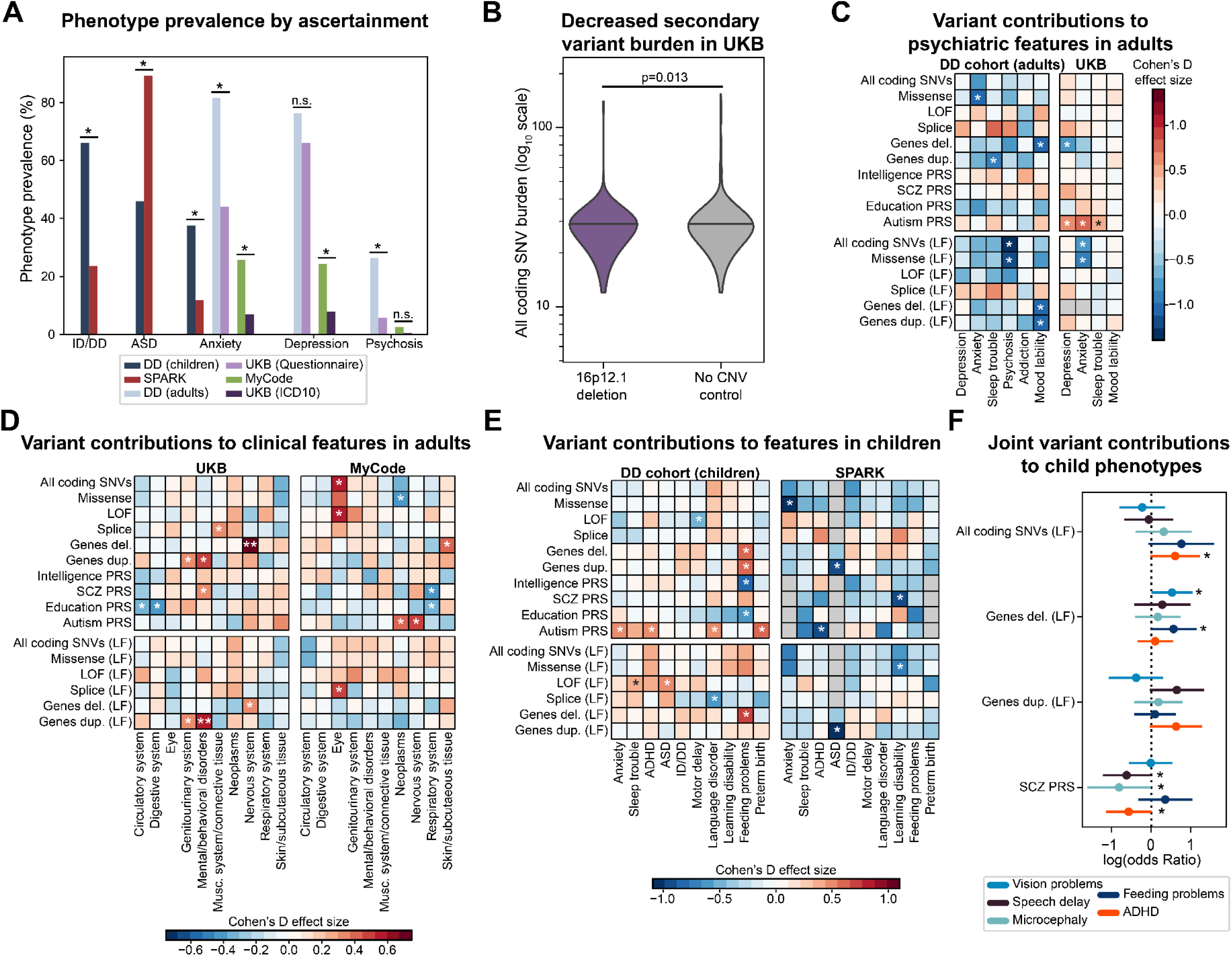
Effects of ascertainment on associations of 16p12.1 deletion. (**A**) Prevalence of phenotypes among adults and children with 16p12.1 deletion from four ascertainments: DD cohort (adults n=38, children n=93-151), SPARK (n=51-56), UK Biobank (UKB; questionnaire n=50-53, ICD10 n=217), and MyCode (n=160). Fisher’s exact test, *p≤0.05. **(B)** Distribution of rare secondary SNVs in UKB individuals with 16p12.1 deletion (n=240, left) and age and sex-matched controls without large rare (>500kb) CNVs (n=2,640, right). P-value from two-tailed t-test. **(C)** Associations of secondary variant burden with select psychiatric phenotypes derived from clinical questionnaires in 16p12.1 deletion adults from DD cohort (n=24-31) and UKB (n=46-249). Two-tailed t-test, *p<0.05. **(D)** Associations of secondary variant burden with select clinical phenotypes derived from EHR data (ICD10 codes) in 16p12.1 deletion individuals from UKB (n=187-218) and MyCode (n=143-159). Two-tailed t-test, *p≤0.05. **Benjamini-Hochberg FDR≤0.05. **(E)** Associations of secondary variant burden and select developmental phenotypes in children with 16p12.1 deletion from the DD cohort (n=67-125) and SPARK (n=27-56). Two-tailed t-test, *p≤0.05. **(F)** Associations of secondary variant burden and developmental phenotypes from joint logistic models of 16p12.1 deletion children from the DD and SPARK cohorts (n=98-125). Joint models for non-LF are shown in **Fig. S4J**, and joint models for adults are shown in **Fig. S4H-I**. *p≤0.05.

We next investigated how patterns of secondary variants differed across cohorts. Adult deletion carriers in UKB showed decreased burden of additional rare coding SNVs compared to individuals without large CNVs (p=0.013) (**Fig. 5B, Table S4C**). Reduced secondary variant burden in 16p12.1 deletion carriers may explain the less severe features observed in the UKB compared to those typically observed among deletion carriers in the clinic. This trend was reversed in SPARK, where individuals with the deletion had an increased burden of SNVs (LF) compared to individuals without large CNVs (p=0.048) (**Fig. S4E, Table S4C**). Thus, we observed a higher rare variant burden in deletion carriers compared to controls in cohorts with more severe disease ascertainment (SPARK) and reduced burden in cohorts with less severe ascertainment (UKB). We also directly compared the variant burden between deletion carriers in the DD cohort to identically processed data from eight deletion carriers in the Estonian Biobank^62^. As expected, Estonian Biobank carriers showed a depletion of missense (FDR≤0.012) and non-coding SNVs (FDR≤0.019) compared with probands and carrier parents in the DD cohort (**Fig. S4F, Table S4J**).

We next assessed how the relationship of secondary variants and phenotypes varies by ascertainment by assessing the burden of variant classes in individuals with and without specific phenotypes across cohorts. We found similar trends for psychiatric features based on self-reported questionnaires for adults in both the DD cohort and UKB (**Fig. 5C, Table S4D**). For example, autism PRS was associated with depression, anxiety, and sleep disturbance (p≤0.037) in UKB (**Fig. 5C, Table S4D**). Conversely, rare variants were negatively associated with psychiatric features in both UKB and DD cohorts, including SNVs (LF) with anxiety in UKB (p=0.010) and psychosis in DD (p=0.003), and deletions with depression in UKB (p=0.016) and mood lability in DD (p=0.027). These data suggest potential opposing roles of PRS and rare variants towards specific psychiatric features. We further compared secondary variant profiles for broader groups of clinical features represented by ICD10 chapters in UKB and MyCode, in contrast to assessment of specific psychiatric features from questionnaires. In UKB, nervous system features were associated with deletions (FDR=0.007), while mental health features were associated with duplications (LF) (FDR=0.035) and schizophrenia PRS (p=0.045) (**Fig. 5D, Table S4E**). In MyCode, eye phenotypes were associated with SNVs (p=0.016), while autism PRS was associated with both nervous system defects and cancer (p≤0.039) (**Fig. 5D, Table S4E**). These differences in genotype-phenotype patterns reflect ascertainment differences between the cohorts, potentially due to healthcare system differences, phenotyping modalities, or biases stemming from healthy volunteers versus clinical patients.

We further observed differences in children with the deletion from the DD cohort and those in SPARK. In general, both rare variants and PRS were associated with increased risk for neurodevelopmental features in DD probands (for example, duplications and deletions for feeding difficulty; p≤0.028) but decreased risk in SPARK probands (for example, decreased missense variants in individuals with anxiety; p=0.025) (**Fig. 5E, Table S4F**). In fact, individuals with ADHD had increased autism PRS in the DD cohort (p=0.017) but decreased autism PRS risk in SPARK (p=0.035) (**Fig. 5E, Table S4F**). This trend reflects the influence of ascertainment towards variant-phenotype associations, where secondary variants may not show the expected associations in highly ascertained cohorts due to saturated genetic risk for the ascertained phenotype, such as ADHD and autism PRS in SPARK (**Fig. S4G**). Overall, we found marked differences in secondary variant-phenotype associations between cohorts (**Fig. 6C-E**), which potentially explains the variable phenotypic trajectories of the deletion observed across ascertainments.

**Figure 6.**
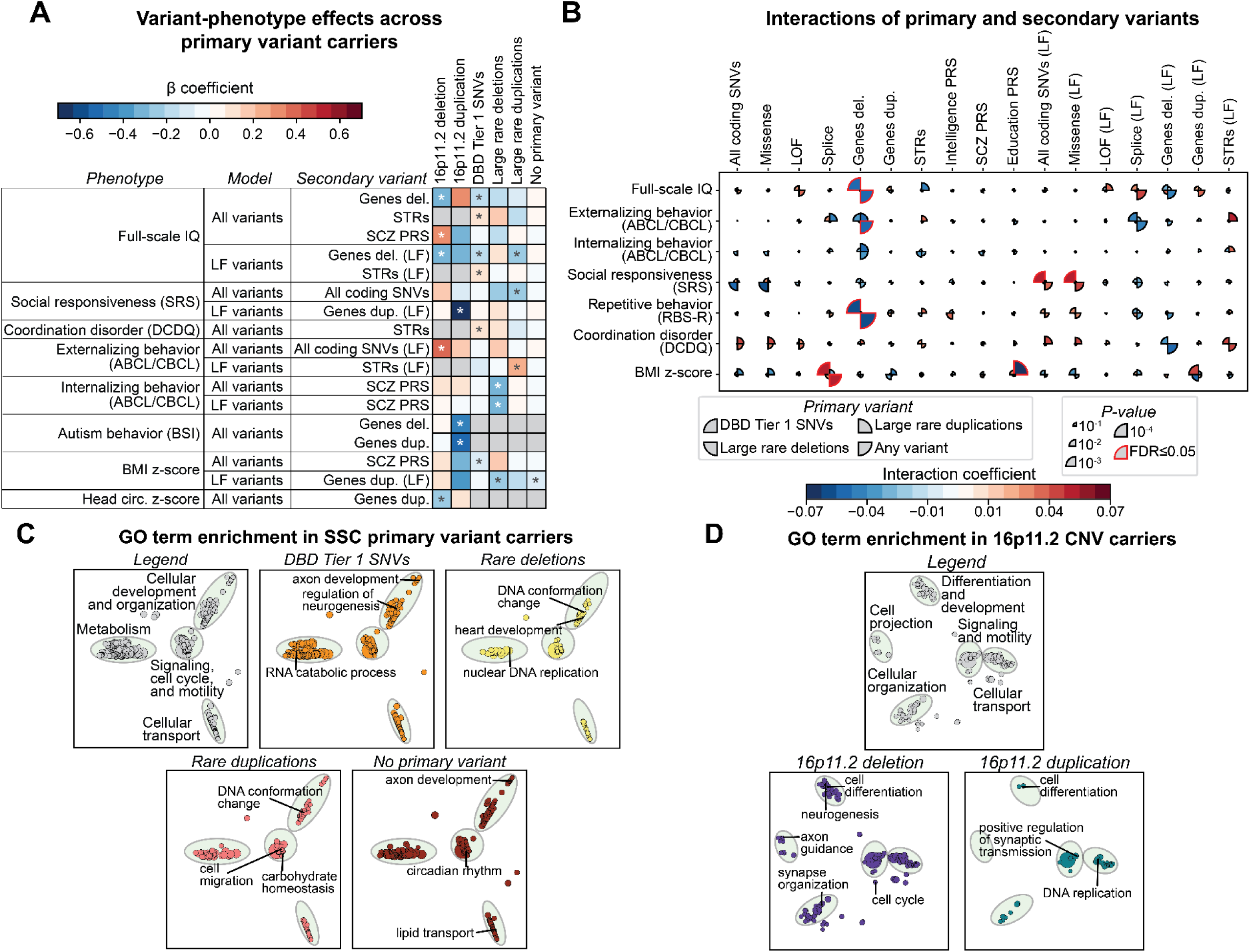
Secondary variant associations in probands with primary variants. (**A**) Heatmap shows β coefficients from select linear regression models for secondary variant burden (y-axis, third column) towards quantitative developmental phenotypes (y-axis, first column) in probands from SSC and Simons Searchlight cohorts with different classes of primary variants (x-axis) (n=21-660). *p≤0.05. **(B)** β coefficients from linear regression models examining interactions between primary variants (pie chart slices) and specific secondary variant classes (x-axis) towards quantitative phenotypes (y-axis) in SSC probands (n=1,597-2,591). Color of pie chart slices indicate interaction coefficients, and size of pie chart slices indicate p-value for strength of interaction coefficient. Red highlights indicate Benjamini-Hochberg FDR ≤0.05 **(C-D)** Gene Ontology (GO) biological process terms enriched among secondary variants observed in **(C)** probands with different classes of primary variants from the SSC cohort and **(D)** probands with 16p11.2 deletions and duplications from the Searchlight cohort. Circles represent individual GO terms, clustered based on semantic similarity into broad categories (green ovals, as defined in the two “legend” plots). Size of each circle represents the number of genes in each term, such that broader terms are larger. Colored circles in each plot indicate significant enrichment of the GO terms for the given primary variant.

We finally combined individuals with the 16p12.1 deletion across cohorts and developed logistic regression models to identify variant-phenotype associations independent of ascertainment bias. We found 12 associations among children in the combined SPARK and DD cohorts (n=84-125), including SNVs with preterm birth (logOR=1.24, p=0.039), deletions with vision problems (logOR=0.878, p=0.018), and SNVs (LF) with ADHD (logOR=0.602, p=0.049) (**Fig. 5F**, **Fig. S4J**, **Table S4G**). Across adults in the DD, UKB, and MyCode cohorts (n=331), duplications were associated with anxiety (logOR=0.092, p=0.004) (**Fig. S4H, Table S4G**).

When examining EHR-derived features in UKB and MyCode (n=321), duplications were associated with circulatory system features (logOR=0.278, p=0.037) and deletions were associated with nervous system (logOR=0.352, p=0.016) and skin/subcutaneous tissue (logOR=0.336, p=0.020) phenotypes (**Fig. S4I**, **Table S4G**). Thus, leveraging combined data from multiple cohorts allowed for the increased statistical power necessary for deriving robust variant-phenotype associations across ascertainments.

### Differing contributions of secondary variants by primary variant ascertainment

To extend our findings beyond the 16p12.1 deletion, we assessed contributions of secondary variants towards developmental, cognitive, and behavioral features among 1,479 probands with different rare pathogenic CNVs or SNVs in known neurodevelopmental genes who were ascertained for the same disorder, i.e. autism (**Fig. 1**). We first assessed 128 probands with reciprocal 16p11.2 deletions (n=91) and duplications (n=37) in the Simon Searchlight cohort^63^ and found eight variant-phenotype associations using linear regression models (**Fig. 7A**, **Table S6A**). Among 16p11.2 deletion probands, schizophrenia PRS contributed to higher full-scale IQ (ß=0.343, p=0.020), while secondary deletions contributed to decreased IQ (ß=-0.283, p=0.040), similar to previous findings^11^ (**Fig. 6A, Fig. S5A, Table S5A**). Different trends were observed in 16p11.2 duplication probands; deletions and duplications were negatively associated with autism behavioral features (BSI; duplications: ß=-0.497, p=0.022; deletions: ß=-0.432, p=0.037) and duplications (LF) were negatively associated with SRS (ß=-0.701, p=0.002) (**Fig. 6A, Fig. S5A, Table S5A)**. Orthogonal correlation analyses revealed several other trends, including opposing effects of secondary duplications towards BSI (16p11.2 deletion individuals: r=0.241, p=0.023; 16p11.2 duplication individuals: r=-0.391, p=0.019) (**Fig. S5B, Table S5E**).

We next assessed 214 probands with a more heterogeneous set of large CNVs, including pathogenic deletions and duplications^33^ at 15q13.3, 3q29, and 16p13.11, from SSC^64^. Among probands with large deletions, linear regression models uncovered negative associations between secondary duplications (LF) with BMI (ß =-0.275, p=0.049), while secondary deletions (LF) were associated with decreased IQ in probands with large duplications (ß =-0.255, p=0.021) (**Fig. 6A, Fig. S6A, Table S5A)**. Correlation analyses revealed additional associations, including SNVs with coordination impairment in probands with large duplications (DCDQ, r=0.178, p=0.042) and decreased SRS in those with large deletions (r=-0.246, p=0.035) (**Fig. S6B, Table S5F**). We further assessed 1,237 SSC probands with SNVs disrupting canonical neurodevelopmental genes^44^, such as *CHD8*, *DYRK1A*, *SCN1A*, and *PTEN*. We again identified a negative association for deletions (LF) with IQ (ß =-0.154, p=5.23×10^−5^), while STRs were associated with increased IQ (ß=0.093, p=0.015) (**Fig. 6A, Fig. S6A, Table S5A**). Correlation analyses uncovered additional effects, such as externalizing behavior (r=-0.111, p=0.001) and repetitive behavior (r=-0.083, p=0.017) negatively correlating with educational attainment PRS (**Fig. S6B, Table S5F**). Thus, we found some consistent patterns across primary variant ascertainments, such as negative effects of rare deletions on IQ, while secondary variant effects on other features were more dependent on the primary variant context.

We additionally examined secondary variants in 1,084 SSC probands without primary variants in the above categories to assess the role of the genetic background outside of a primary variant context. The only observed association from regression analysis was for duplications (LF) and lower BMI (ß=-0.087, p=0.031) (**Fig. 6A, Table S5A**). The paucity of associations in the absence of primary variants suggests that secondary variant classes mostly exert their effects through interactions with primary variants instead of contributing directly towards disease phenotypes. To assess this, we used linear models to identify interactions between primary and secondary variants towards clinical features. We found ten instances of multiplicative interactions, including primary SNVs and secondary SNVs (LF) towards SRS (ß=0.054, FDR=0.034) as well as primary SNVs and secondary deletions towards full-scale IQ (ß=-0.058, FDR=0.020) and repetitive behavior (ß=-0.062, FDR=0.011) (**Fig. 6B**, **Table S5B**). Notably, these interactions tended to be primary variant-specific, further supporting the hypothesis that secondary variant effects are influenced by primary variant context.

The relevance of primary variant context was further evident when we assessed the biological functions of secondary variants (**Fig. 6C-D)**. For example, secondary variants in probands with primary SNVs showed specific enrichments for neuronal development and cell-to-cell signaling, while probands with primary deletions showed enrichments for DNA repair and replication (**Table S5C**). Secondary variants in 16p11.2 duplication probands were enriched for DNA replication and synaptic transmission regulation, while variants in probands with the reciprocal deletion were enriched for cell cycle regulation and synapse organization (**Table S5D**). In fact, multiple genes within the 16p11.2 deletion have similar molecular functions (i.e. *MAPK3* and cell cycle regulation^65^), and many of the same GO enrichments, including neuronal differentiation and projection, were identified among differentially expressed genes in animal models of genes within the 16p11.2 region^66^ (**Table S5D**). We therefore posit that modifier variants influence developmental features by acting additively or synergistically in molecular pathways disrupted by the primary variant, further underscoring the importance of primary variant context^67^.

## DISCUSSION

Our comprehensive analysis of 2,252 individuals with primary variants from several diverse cohorts allowed us to find strong evidence that modifier variants confer distinct risks towards different developmental and clinical phenotypes. These effects are contingent upon the context of the primary variant, secondary variant class and function, phenotype of interest, and cohort ascertainment. Our results emphasize the importance of assessing a full spectrum of genomic variants in a variety of contexts to unravel the etiology of heterogeneous clinical features observed in individuals with the same primary variants.

Several of our results expand on previous work to identify mechanisms for variable expressivity of pathogenic variants and may help refine the broader roles of modifier variants in complex disorders. First, we identified roles for a wide set of rare variants towards developmental features of the 16p12.1 deletion, including noncoding variants and STRs. These findings expand previous definitions of secondary variants beyond additional CNVs or rare coding SNVs^28,33^. Second, we expanded the role for PRS towards various developmental and psychiatric phenotypes in individuals with pathogenic CNVs. These findings are in line with recent studies that have identified roles for PRS as modifiers of specific phenotypes of pathogenic CNVs, such as BMI z-scores for 16p11.2 CNVs^10^. Third, we observed cases of compounding variant burden across generations of 16p12.1 deletion carriers, which could explain our previous findings correlating rare variant burden with family history of psychiatric disorders^28,54^. This phenomenon could be attributed to assortative mating on psychiatric features between deletion and non-deletion parents; in fact, we recently reported that 16p12.1 deletion spouse pairs show strong correlations for psychiatric disorders, which may lead to increasing genetic risk over generations^68^. Fourth, while disease-relevant secondary variants contributed to multiple genetic diagnoses, we did not find any instance where a single variant solely accounted for all phenotypes observed in a proband, suggesting that secondary variants modify effects of the deletion.

Ascertainment bias can preclude a more thorough assessment of a full spectrum of phenotypes due to primary variants, including subtler or progressive effects, as deeper evaluations are typically restricted to individuals with specific disorders^69,70^. We therefore examined the effects of cohort ascertainment within a single primary variant-specific context. While the 16p12.1 deletion contributed to clinical outcomes across disease-ascertained and general population cohorts, the specific phenotypic trajectories and the influence of secondary variants both differed substantially across ascertainments. Our findings have several implications, as management of primary variant-related symptoms may differ in individuals evaluated for severe developmental features versus those with other medical features, where the variant may first present as an incidental finding^19^. Thus, a shift in treatment focus from just the primary variant to all variants in an affected individual could allow for more effective management of individuals who carry primary variants.

The observed variability among secondary variant-phenotype patterns in each cohort makes it difficult to identify consistent patterns across ascertainments, limiting the generalizability of variant association studies conducted in a single cohort. In fact, the only consistent trends we observed across primary variants, such as 16p11.2 deletion and rare disease-associated SNVs, were for reduced IQ correlating with increased rare secondary variants, mirroring previous studies^11^. Joint models that integrate data across cohorts with appropriate covariates can be used to overcome this ascertainment bias; for example, we found several significant associations using joint models of 16p12.1 deletion carriers, including rare SNV burden towards ADHD and speech delay in affected children. More broadly across primary variant and ascertainments, we observed general trends for more PRS effects towards psychiatric features, such as autism PRS and ADHD in DD children and SCZ PRS for mental/behavioral disorders in UKB, and more rare variant effects towards cognitive features, such as rare deletions (LF) with full-scale IQ in SSC probands with disease-associated SNVs and 16p11.2 deletion probands. Both trends mirror previous variant-phenotype associations in individuals with autism outside of a primary variant context^71^. However, some exceptions to these patterns exist: SSC probands with rare duplications and 16p11.2 duplication probands show higher effects of rare secondary variants towards psychiatric phenotypes, including negative associations of splice variants with externalizing behavior in both groups. Further, 16p12.1 deletion carriers from SPARK and 16p11.2 deletion probands show greater effects of PRS towards cognitive features, such as the positive association of SCZ PRS and full-scale IQ in 16p11.2 deletion probands and a negative association of SCZ PRS with learning disability in SPARK, potentially a facet of autism-specific genetic etiology for cognitive features. Therefore, when describing genotype-phenotype associations in a primary variant context, future studies should strive to assess multiple independent cohorts with different ascertainments to determine the extent that ascertainment could bias their results.

Despite assessing the contributions of multiple secondary variant classes towards specific clinical features of pathogenic variants, much of the genetic etiology for these features is still not accounted for in our study. Several factors could account for the unexplained variance, including environmental factors or additional variant classes such as inversions, as well as those that could explain ascertainment variability, including population-specific effects in the genetic background. Another under-studied source of the unexplained variance could be non-additive interactions between variants. Only a small number of synergistic variant interactions have been identified to date in complex genomic disorders^72^, and very large cohorts will be required to quantify the effects of these interactions towards clinical features^73^. Here, we identified non-additive effects of primary and secondary variants among children ascertained for autism. Molecular studies could help unravel the mechanisms by which modifier variants interact with primary variants to influence their phenotypes. For instance, we previously found 11 cases where secondary variants synergistically altered the expression of genes dysregulated by the deletion in patient-derived LCL models^54^. While the overall effects of such interactions will likely explain only a portion of the unexplained variance, they may play an outsized role in CNV disorders due to the potential for interactions among multiple genes within the primary variant^26^.

Overall, we identified family-, phenotype-, ascertainment-, and primary variant-specific patterns of secondary variants that influence the variable expressivity of the 16p12.1 deletion and other primary variants. Our study emphasizes the complexity of neurodevelopmental disorders even after a causal variant is identified, suggestive of an oligogenic model for disease pathogenicity^74^. For researchers and clinicians alike, our study highlights the importance of understanding the influence of cohort ascertainment and thoroughly investigating genomic variants with smaller effect sizes. The complexity of the 16p12.1 deletion and other genomic disorders calls for personalized medicine approaches that fully account for individual-level phenotypic presentation, family history, and genome-wide variant profile towards counseling, management, and potential treatment.

### Limitations of this study

One limitation of our study is the relatively low sample size of families with the 16p12.1 deletion and other primary variants. While this study represents one of the largest cohorts of individuals with the same pathogenic variant to date and is well-powered for assessing changes in rare coding SNV burden among deletion carriers, the study is under-powered for identifying enrichments of individual variants or genes towards specific phenotypes. Additionally, while our study captures major themes regarding variable expressivity of pathogenic variants, some specific associations have only nominal significance. Larger cohorts will allow for further study of these trends and could uncover roles for specific genes or molecular pathways towards clinical features. Finally, while we were able to leverage data from 773 16p12.1 deletion individuals from multiple ascertainments, differences in genotyping and phenotyping methods precluded direct comparisons between cohorts.

## AUTHOR CONTRIBUTIONS

M.J., C.S., A.T., L.P., and S.G. designed the study and analyses. C.S., L.P., E.H., and L.R. recruited patients and conducted interviews, harmonized phenotypic data from interviews and medical records, and extracted DNA from blood for WGS sequencing. C.T. and C.L.M. assisted with collection and analysis of quantitative phenotypic data. Other authors provided de-identified DNA, blood samples, or genomic and phenotypic data of 16p12.1 deletion families to the study. M.J., C.S., A.T., D.B., and V.K.P. designed bioinformatics pipelines to identify variants, uniformly processed sequencing data from 16p12.1 deletion and external cohorts, and performed all statistical, enrichment, and modeling analysis. V.R. and H.S. assisted with design of the PRS calculations and modeling approaches. M.J., C.S., A.T., L.P, and S.G. wrote the manuscript with approval from all authors.

## Data Availability

Whole genome sequencing and SNP microarray data generated in this study are available at NCBI dbGaP phs002450.v2.p1. Statistical analyses and experimental results for the data presented in Figs. 2-7 and associated supplementary figures are available in Tables S2-S6. 

https://www.ncbi.nlm.nih.gov/projects/gap/cgi-bin/study.cgi?study_id=phs002450.v2.p1

https://github.com/girirajanlab/16p12_WGS_project

## ACKNOWLEDGEMENTS

This work was supported by NIH R01-GM121907 and resources from the Huck Institutes of the Life Sciences to S.G. M.J. and C.S. were supported by NIH T32-GM102057. A.T. was supported by NIH T32-LM012415. L.P. was supported by Fulbright Commission Uruguay-ANII. A.R. was supported by grants from the Swiss National Science Foundation 31003A_182632. S.B. was supported by the NIHR Manchester Biomedical Research Centre (NIHR203308). We thank Craig Praul and the Penn State Genomics Core Facility for assistance with designing the WGS sequencing strategy, Abby Hare-Harris (Geisinger ADMI) for assistance with RedCap analysis of quantitative phenotypic data, Veera Rajagopal (Regenron Pharmaceuticals) and Bertrand Isidor (CHU Nantes) for useful comments on the manuscript, and Jianyu Yang, Sarah Dwiekat, and Edmundo Torres-Rodriguez (Penn State) for assistance with curating annotation data for variant analysis. We are grateful to all of the families in each cohort (DD, SPARK, Simons Searchlight, SSC, MyCode, and UK Biobank) as well as clinical sites and staff who participated in the study. We thank the SSC principal investigators (A. Beaudet, R. Bernier, J. Constantino, E. Cook, E. Fombonne, D. Geschwind, R. Goin-Kochel, E. Hanson, D. Grice, A. Klin, D. Ledbetter, C. Lord, C. Martin, D. Martin, R. Maxim, J. Miles, O. Ousley, K. Pelphrey, B. Peterson, J. Piggot, C. Saulnier, M. State, W. Stone, J. Sutcliffe, C. Walsh, Z. Warren, E. Wijsman) as well as the Simons Searchlight Consortium. We appreciate obtaining access to genomic and phenotypic data on SFARI Base. Approved researchers can obtain the SPARK, SSC, and Simons Searchlight population data sets described in this study by applying at https://www.base.sfari.org. This research has been conducted using data from UK Biobank. More information about UK Biobank is available at https://www.ukbiobank.ac.uk/.

## DECLARATION OF INTERESTS

The authors declare no competing interests.

## METHODS

### 16p12.1#deletion developmental delay cohort description

We analyzed genomic and phenotypic data from a cohort of 452 individuals belonging to 132 families with the 16p12.1 deletion, which we refer to as the Developmental Delay cohort (“DD cohort”). (**Fig. 1, Table S1A**). These families comprised single probands (n=14), parent-child pairs (n=13), complete trios (n=97), and extended families, including eight families with three generations and 22 with multiple affected children. The deletion was identified through prior clinical diagnostic tests for ID/DD or other developmental disorders in probands. We note that 10 individuals from eight families with the 16p12.1 deletion representing an unselected population from the Estonian BioBank were included as a comparison group within the DD cohort^62^. Whole genome sequencing was performed on 287 individuals (107 probands), including the eight Estonian BioBank families, and microarray experiments were performed for 368 individuals. Informed consent was obtained from families recruited directly according to a protocol approved by the Pennsylvania State University Institutional Review Board (IRB #STUDY00000278), while de-identified information was obtained from families recruited through clinics according to another approved protocol (IRB #STUDY00017269). A list of all individuals in the DD cohort, including familial membership and 16p12.1 deletion status, is available in **Table S1A**. Note that some information that could be construed as personally identifiable is summarized in **Table S1A** (i.e., age ranges instead of age values); full datasets are available upon request from the authors for purposes of reproducibility (i.e., for the model covariates). We also note that sample and family identifiers used in **Table S1A** are specific for this study and were not known outside of the research group. Power calculations for detecting changes in rare variant burden within deletion family members (**Fig. S1B**) were based on estimated effect sizes of burden differences between 16p12.1 deletion probands and parents (coding SNVs and CNVs) or between autism probands and parents (non-coding SNVs and STRs) from previous studies^28,75,76^.

### Phenotypic analysis

Individual-level phenotypic data described below (phenotypic domain complexity scores, quantitate phenotypes, and family history status) are available in **Table S1A**.

#### (a)#Collection and analysis of clinical features

We collected detailed medical history and used clinician-, guardian-(for children), or self-(for adults) reported standardized questionnaires to assess developmental phenotypes in children (average age=10.1 years) and psychiatric features in adults. Questionnaires for children assessed neuropsychiatric and developmental features, anthropomorphic measures, congenital abnormalities in multiple organ systems, and family history of medical or psychiatric disorders. Phone surveys were conducted to fill in missing information, and families were recontacted every 3-4 years to track longitudinal data and to note any later-onset clinical features. We analyzed phenotypes for each individual by calculating “complexity scores” for clinical features within specific domains, as well as measured specific quantitative features (see “Assessment of quantitative phenotypes” below).

*For children*, we first grouped clinical features into six broadly defined phenotypic domains: (i) ID/DD, (ii) behavioral phenotypes, (iii) psychiatric features, (iv) nervous system defects, (v) craniofacial and skeletal abnormalities, and (vi) congenital abnormalities (**Fig. 2A, Table S1A-B**). We determined complexity scores ranging from 0 to 4 or 5 for each phenotypic domain by assessing the total number of affected phenotypic sub-domains in each child. The full list of phenotypes considered for each sub-domain is available in **Table S1B**. Presence of at least one clinical feature within a sub-domain added an additional point of complexity to the total score, but additional phenotypes within the same sub-domain added no additional complexity score. For example, proband P1C_001 had three nervous system-related phenotypes (tremors, abnormal gait, and abnormal brain morphology) grouped into two sub-domains (nervous system abnormalities and nervous system morphology defects), and therefore received two points for complexity. We note that younger probands were not assessed for psychiatric domains based on the typical age of onset, such as for schizophrenia (**Table S1C**). As most probands (92%) exhibited >1 feature within the ID/DD domain, we focused on the other five phenotypic domains for downstream analysis.

*For adult family members*, we binned and calculated complexity scores in a similar manner for clinical features within four domains, including (i) cognitive (ID, learning difficulty), (ii) psychiatric (schizophrenia, bipolar disorder), (iii) depression/anxiety, and (iv) addiction phenotypes (**Fig. 2B**). We note that most phenotypes for early-onset behavioral and developmental features assessed in children were not examined for adult family members.

#### (b)#Assessment of quantitative phenotypes

We performed online quantitative assessments using the Hansen Research Services Matrix Adaptive Test (HRS-MAT) for non-verbal IQ^34^ and Social Responsiveness Scale (SRS) for autism-related social behavior^35^ (**Fig. 2C**). HRS-MAT was self-administered to participants through an online platform, while the SRS was administered through a RedCap-based survey platform maintained by the Geisinger Autism and Developmental Medicine Institute. The SRS assessment was self-reported if the participant was over 18 years or completed by parents or guardians for children under the age of 18 years. Body Mass Index (BMI) and head circumference were obtained from medical records or self/guardian-reports or, for BMI, calculated from height and weight data obtained from medical records or self/guardian-reports. Both BMI and head circumference were converted into age– and sex-adjusted z-scores^77,78^. We further obtained SRS, BMI, and head circumference z-scores for probands in the SRS and Simons Searchlight cohorts (see below), while the mean HRS-MAT score in SSC probands was obtained from Hansen, 2019 ^34^. Differences in phenotype distributions between groups of 16p12.1 deletion family members and between sets of probands with different primary variants were calculated using one– and two-tailed Mann Whitney tests, respectively (**Table S2A**). Differences of proband scores from a defined mean were calculated using one-tailed one-sample t-tests (**Table S2A**).

#### (c)#Developmental milestones

We assessed the achievement of developmental milestones in children from the DD cohort based on CDC guidelines^36^. Parents/guardians of children reported the ages at which children achieved 12 milestones, including age first smiled, rolled over, crawled, walked, and spoke. Age of milestone attainment for all available samples is reported in **Table S1A**. Differences in milestone achievement between probands and their siblings/cousins were assessed using one-tailed t-tests (**Table S2B**).

### DNA extraction and whole-genome sequencing

We performed DNA extraction and whole-genome sequencing on 287 individuals in the DD cohort (**Table S1A)**. Genomic DNA was extracted from peripheral blood samples from some participants using the QIAamp DNA Blood Maxi extraction kit (Qiagen, Hilden, Germany) while clinical collaborators sent isolated DNA from other participants. Illumina TruSeq DNA PCR-free libraries (San Diego, CA, USA) were constructed for 150bp paired-end whole-genome sequencing using Illumina HiSeq X by Macrogen Labs (Rockville, MD, USA). Samples were sequenced at an average 35.7X coverage, or 716.2 M reads/sample, with 94.9% of reads mapping to the human genome. After processing for quality control using Trimmomatic^79^ (leading:5, trailing:5, and slidingwindow:4:20 parameters), sequences were aligned to the hg19 reference genome using BWA v.0.7.13^80^, and sorted and indexed using Samtools v.1.9^81^. We note that sequencing data from 163 individuals was described previously^54,68^.

We used SNP microarrays for copy-number variant validation and genotyping experiments (i.e. CNV calling and polygenic risk score calculation) for 368 individuals. Samples were run on Illumina OmniExpress 24 v.1.1 microarrays at Northwest Genomics Center in the University of Washington (Seattle, WA, USA). We note that microarray data of 208 individuals was described previously^28,54,68^.

### Identification and annotation of single-nucleotide variants

We identified SNVs and small indels using the GATK Best Practices pipeline^82^, followed by quality control and extensive variant and gene-level annotations. Duplicate removal with PicardTools was followed by base-pair quality score recalibration and variant calling for each sample using GATK HaplotypeCaller v.3.8. We then merged calls from all samples using GATK GenotypeGVCFs v.4.0.11, performed variant quality score recalibration to finalize variant calls, and used Vcfanno to annotate variants with GnomAD frequency^83,84^. All calls were filtered for QUAL ≥50, allele balance between 0.25 and 0.75 or ≥0.9, read depth ≥8, QUAL/alternative read depth ≥1.5, GnomAD frequency ≤0.1% (or not present), and intracohort frequency ≤10 to account for technical differences between our data and GnomAD. We annotated coding and noncoding variants within genes from GENCODE^85^ v19 using ANNOVAR^86^ and Vcfanno^83^ as follows: *(a) Coding SNVs*: Rare coding variants were filtered for loss-of-function (LOF), missense, or splicing exonic variants in protein-coding genes, and annotated with CADD Phred-like scores, presence in ClinVar database, the gene-level pathogenicity metric LOEUF, and gene-level phenotype associations using OMIM^37,42,43,87^. Missense and splice variants were filtered to include only those with a CADD score ≥25. *(b) Noncoding SNVs*: All rare variants located 1kbp upstream of a gene transcription start site were classified as promoter variants, while genes within the 5’ UTR were classified as 5’ UTR variants. Fetal brain-active enhancer regions were identified using chromatin state data from the Roadmap Epigenomics consortium^88^ (states 6, 7, and 12 in the fetal brain), and rare variants in those regions were classified as enhancer variants. Rare SNV burden for all available individuals are listed in **Table S1A**.

### Identification of copy-number variants and short tandem repeats

CNVs were called from both microarray data using PennCNV^89^ and WGS data using CNVnator v.0.4.1, Lumpy-sv v.0.2.13 with Smoove v.0.2.5, Delly v.1, and Manta v.1.6.0^90–93^. For CNVs >50 kbp in length, we used a union of PennCNV and CNVnator calls. For CNVs <50 kbp, we used CNVs called by least two of CNVnator, Lumpy, Manta, or Delly, defined by 50% reciprocal overlap. All CNVs were annotated for 50% reciprocal overlap with known pathogenic CNVs^33^. WGS-based CNV calls were filtered to remove calls with >50% overlap with centromeres, segmental duplications, regions of low mappability, and V(D)J recombination regions, while microarray CNVs were filtered to remove samples with >50% overlap with centromeres, telomeres, and segmental duplications^94^. Known pathogenic CNVs were excluded from this filter^33^. All CNVs were then filtered for GnomAD-SV frequency^95^ <0.1% and intracohort frequency ≤10, and >50 kb CNVs were additionally filtered for <0.1% frequency in a control cohort^96^. All CNVs were finally filtered for those intersecting at least one protein-coding gene, using gene annotations from GENCODE v19^85^.

We identified STR expansions from WGS data using the GangSTR v.2.4 (reference file v.13.1) and TRTools pipelines^97,98^. We filtered calls with read depth >20 and <1000, excluding reads that were not spanning and bounding the STR locus and calls with maximum likelihood estimates not within the confidence interval using dumpSTR. After merging the calls with mergeSTR, we ran dumpSTR with population level filters, including locus call rate >0.8 and departure from Hardy Weinberg equilibrium (Fisher’s exact p-value) >0.00001, and removed loci that overlapped with segmental duplications. For chromosome X, the Hardy-Weinberg equilibrium p-value was calculated from female samples only. For each family, we extracted the STR loci that passed variant filtering, and used GangSTR v2.5 to call STR variants, which were used in subsequent analyses. We defined STR expansions as STR variants with lengths >2SD higher than the average of STR lengths among all individuals in our in-house cohort of individuals with WGS data at a particular locus. STR expansions spanning protein coding regions defined by GENCODE v19^85^ were selected for downstream analysis. All CNV and STR genes were further annotated for pathogenicity metrics (LOEUF score^37^). The number of genes affected by rare CNVs and the number of STR expansions for all individuals with available WGS data are listed in **Table S1A**.

### Polygenic risk score calculations

Using microarray data, we calculated polygenic risk scores for educational attainment^38^, intelligence^39^, schizophrenia^40^, and autism^41^ among the samples in the DD cohort, based on standardized bioinformatics pipelines for quality control^99^. We first downloaded summary statistics from four recent GWAS studies of neuropsychiatric traits, and filtered SNPs for imputation INFO scores >0.8 and removed duplicate and ambiguous SNPs. We then merged SNP genotype data from different microarray batches together using PLINK. Initial quality control removed SNPs with minor allele frequency <0.05, Hardy-Weinberg equilibrium <1.0×10^−6^, and genotype rate <0.01, along with samples missing >1% of genotypes. We used the HRC-1000 Genomes Imputation toolkit (https://www.well.ox.ac.uk/~wrayner/tools) to process PLINK files into individual chromosomes for imputation, and VcfCooker (https://genome.sph.umich.edu/wiki/VcfCooker) to convert PLINK files to VCF files. Microarray-based SNPs were imputed using the TOPMed v.r2 imputation server using Eagle v2.4 for phasing^100^. After imputation, VCF files were converted back to PLINK format, and SNPs were again filtered with identical QC filters. Additional QC filters included removing samples with >±3SD of the mean heterozygosity rate. We also selected individuals with European ancestry, based on imputed genetic ancestry (calculated using Peddy v.0.4.8 with 1000 Genomes-based population panel) or self-reported ancestry, for downstream analysis^101^. Finally, we performed strand-flipping to match SNPs in microarray data with the GWAS summary statistics. To calculate PRS, we used standardized pipelines for the LDPred2 software package, which uses Bayesian approaches to optimize parameters for PRS calculation^102^. Briefly, we filtered the four sets of GWAS summary statistics for SNPs present in the HapMap3 dataset^103^, and used 1000 Genomes datasets to calculate linkage disequilibrium matrices for the SNPs^99^. After regressing betas or odds ratios of GWAS SNPs according to linkage disequilibrium, we used the LDPred2-auto model to calculate the four PRS values for all samples with available genotype data. PRS for all available individuals are listed in **Table S1A**.

### Genotype-phenotype statistical and modeling analysis

We performed multiple analyses to compare effects of rare variants and PRS towards different phenotypic domains among 16p12.1 deletion probands or between probands and their carrier and non-carrier parents. When assessing variant effects towards phenotypic domains, we binned probands with different complexity scores for each phenotypic domain into binary groups of roughly equal sizes (i.e., probands with higher versus lower complexity scores) for logistic regression models and burden tests. Burden analysis (paired and independent T-tests) and Pearson’s correlation analyses were calculated in Python v.3.7 using the scipy v1.13.1 *ttest_rel, ttest_ind,* or *pearsonr* functions, respectively. We note that paired t-tests for PRS burden between probands and parents were two-tailed, due to the dual directionality of PRS for different phenotypes, while other t-tests were one-tailed. Benjamini-Hochberg multiple testing correction was performed using the scipy v.1.13.1 *false_discovery_control* function for all statistical analyses unless otherwise stated. Multiple testing was performed separately for analyses with all sets of rare variants and variants filtered for evolutionary constraint (defined by LOEUF<0.35, which are intolerant to loss-of-function variants in the general population^37^; referred to as “(LF)”). FDR values reported in the text are corrected for multiple testing, while p-values are not corrected for or did not pass multiple testing. Sample sizes, test statistics, and corrected and uncorrected p-values for all analyses are available in **Tables S2-S5**.

Logistic and linear regression models for phenotypic variation among probands were performed using the *Logit* and *OLM* functions in statsmodels v.0.14.2, respectively (**Table S3A**). For joint variant regression models, we used three different sets of genetic input variables to test for effects towards phenotypes: (a) all rare variants (sum of SNV, STR, and CNV gene burden) and schizophrenia PRS; (b) SNVs, STRs, duplications, and deletions; and (c) SNVs, STRs, duplications, and deletions restricted to genes with LOEUF scores<0.35 (referred to in the figures as “LF model”). Single variant regression models used only a single variant class as input. All models also included sex as a covariate, while models *b* and *c* also included schizophrenia PRS as a covariate. Additional covariates, such as age and genotype PCs, were not included due to concerns regarding potential overfitting of models with lower sample sizes. The variance explained by the models (R^2^ for linear models and McFadden’s pseudo-R^2^ for logistic models) was calculated for all models. Sample sizes, odds ratios, uncorrected p-values, confidence intervals, and variance statistics for all regression models used in the DD cohort are available in **Tables S3A**.

### Variant enrichment and pathogenicity analysis

#### Gene set enrichment

We assessed enrichment of genes with secondary variants among sets of neurodevelopmental disease genes and genes with neuronal function from several previously published resources^37,44,45,48,104–106^. We identified enrichment of variants in these gene sets by performing Fisher’s Exact tests against the whole genome for each gene list and calculated odds ratios and p-values for genes with variants in the DD cohort using the *contingency.odds_ratio* function from scipy v.1.13.1 (**Table S2G**).

#### Gene ontology

Gene ontology (GO) term enrichment was performed using the Panther API and the GO-Slim Biological Process annotation dataset^107^ (**Table S3B**). The GO term network figures were created using GO term semantic similarity calculated from rrvgo^108^ v1.10.0 in R v4.2.3.

#### Spatio-temporal brain expression

We assessed variant enrichment in genes preferentially expressed in specific brain tissues using the BrainSpan Atlas^52^ and in genes preferentially expressed in specific cell types using single-cell RNA-seq expression data in the M1 motor cortex^53^. We defined preferentially expressed genes as those with expression >2SD higher than the median expression across all tissues or all cell types for that gene. We used Fisher’s exact tests as described above to find the odds that a gene both carries a variant in the DD cohort and is expressed in a specific brain region or cell type (**Table S2E, S2H**).

#### 16p12.1 differentially and co-expressed genes

We used Fisher’s Exact tests as described to calculate enrichment of secondary variants in gene co-expression modules previously identified using WGCNA in lymphoblastoid cell line (LCL) models of the 16p12.1 deletion^54^ (**Table S2F**). We specifically assessed six co-expression modules whose genes showed differential expression between deletion carriers and controls, one of which (black module) also contained two genes within the 16p12.1 deletion region (*POLR3E*, *MOSMO*).

For all enrichments, we applied Benjamini-Hochberg multiple testing correction as described above. Sample sizes, test statistics and corrected and uncorrected p-values for enrichments are listed in **Tables S2** and **S3**.

#### Pathogenic variant analysis

We defined pathogenic SNVs as those that are “Pathogenic” or “Likely pathogenic” in ClinVar for neurodevelopmental phenotypes^42^, or loss-of-function variants in genes that (*a*) have dominant or (if male) X-linked neurodevelopmental OMIM phenotype^43^, (*b*) are a Tier S or Tier 1 SFARI gene, which represent strong candidate autism genes^45^, or (*c*) are in the Tier 1 or Tier 2 gene list from the Developmental Brain Disorder Gene Database, which represent genes with well-documented connections to neurodevelopmental disease^44^ (**Table S1D**). Pathogenic CNVs were identified from 50% reciprocal overlap with a previously published list of CNVs^33^.

### Network analysis

We assessed the connectivity of genes within a previously described brain-specific interaction network. In brief, the network was built using a machine-learning model trained to predict the likelihood of interactions between pairs of genes using brain-specific gene co-expression, protein-protein interaction, and regulatory sequence datasets^49,50^. We restricted the network to genetic interactions with weighted probabilities >2 and calculated the degree for each gene (or number of connections between a particular gene and other genes) as a descriptor of connectivity to other genes in the network. We then binned each gene into one of four quantiles based on the gene’s degree of connectivity and counted the number of times that genes with coding variants fell into each quantile. To calculate empirical p-values, we compared the resulting values to 1000 simulations in which we randomly selected the same number of genes from the genome and counted the number of randomly selected genes that fell into each quantile (**Table S2D**).

### Genotype-phenotype analysis in 16p12.1 deletion samples from other ascertainments

#### (a)#Description of cohorts

We identified individuals carrying 16p12.1 deletions from three additional cohorts, each representing a distinct ascertainment. Individuals in the Simons Powering Autism Research for Knowledge (SPARK) cohort (n=94) were ascertained for families with autism^58^, while the Geisinger MyCode Community Health Initiative (MyCode) (n=160) represents a health care-based cohort^60^ and the UK Biobank^59^ (UKB) (n=250) consists of individuals with a “healthy volunteer” bias^61^. Combined with samples from the DD cohort (after excluding samples with incomplete phenotypic information), we assessed a total of 757 deletion carriers. De-identified data from these cohorts were obtained and analyzed according to a protocol approved by the Pennsylvania State University Institutional Review Board (IRB #STUDY00011008). Data from the UK Biobank was accessed under application 45023. Individuals from the MyCode cohort were recruited during primary care or specialty clinic visits to Geisinger Health System locations, independent of condition, diagnosis, or demographic characteristic. Written informed consent was obtained from adult patients and from the parents or guardians of pediatric patients. The study was conducted with approval from the Geisinger institutional review board.

#### (b)#CNV calling

Carriers of the 16p12.1 deletion in each cohort were identified based on CNV calls from microarray data. Samples from MyCode were genotyped using the Illumina Global Screening Array and Illumina OmniExpressExome-8 Kit. SNP log-r ratio and b-allele frequencies for SPARK samples were downloaded through the SFARI Base portal (https://www.base.sfari.org), while signals for the UK Biobank were accessed through Data Fields 22437 and 22431. CNVs for all cohorts were called using the PennCNV pipeline described above for the DD cohort^89^. Additionally, 2,640 and 356 additional samples without any large (>500kb), rare (<0.1% population frequency) CNVs were identified as controls for additional genetic analysis from the UK Biobank and SPARK, respectively.

#### (c)#SNV calling from sequencing data

SNVs for all three cohorts were identified from whole exome sequencing (WES) data. NimbleGen (SeqCap VCRome) and xGEN probes from Integrated DNA Technologies (IDT) were used for target sequence capture in the MyCode cohort^109,110^. Sequencing was performed by paired end 75bp reads on an Illumina NovaSeq or HiSeq at coverage >20x at >80% of the targeted bases. Alignments and variant calling were based on GRCh38 human genome reference sequence. Variants were called with the WeCall variant caller version 1.1.2 (https://github.com/Genomicsplc/wecall). Whole exome VCFs for SPARK samples were downloaded through the SFARI Base portal (https://www.base.sfari.org). WES VCFs from both cohorts were processed using the same pipeline described above for the DD cohort. WES data from UKB individuals was available as multi-sample project VCFs^111^ in the UK Biobank Research Analysis Platform. After splitting multi-allelic records, we applied the following set of quality control filters using Hail in the DNAnexus platform: (a) variant call rate >90%, (b) Hardy Weinberg equilibrium p-value >10-15, (c) minimum read depth > 10, and (d) at least one sample passing the allelic balance threshold of 0.2. Next, we filtered for variants with an intracohort frequency <0.1% and present in at least two samples. The remaining variants were then annotated using Variant Effect Predictor^112^ (VEP) v.109 and dbNSFP^113^ v.4 to identify their effects on gene transcripts. We specifically annotated variants based on VEP annotations as LOF (transcript ablation, stop gained, frameshift variant, stop lost, and start lost), missense, or splice (splice acceptor variant and splice donor variant). Missense variants were further filtered for those predicted to be deleterious by at least five of nine selected tools (SIFT, LRT, FATHMM, PROVEAN, MetaSVM, MetaLR, PrimateAI, DEOGEN2, and MutationAssessor) available through the dbNSFP database^113^.

#### (c)#Phenotype analysis

We assessed phenotypic data in these cohorts using ICD10 codes derived from electronic health records (MyCode and UKB) and self-reported questionnaire responses (UKB and SPARK) (**Table S4A**). Phenotypic information from SPARK was downloaded from the Simons Foundation through the SFARI Base portal (https://www.base.sfari.org). Electronic health records were available from participants in MyCode^60^. Electronic health records for UKB were identified from Data Fields 41202 and 41204 (main and secondary inpatient ICD10 codes), while questionnaire data was identified from additional Data Fields (**Table S4A**). For harmonization of phenotypic data across cohorts, ICD10 codes were matched to phenotypes from questionnaires, details of which are provided in **Table S4A**.

#### (d)#Secondary variant associations

Comparisons of secondary variant burden between 16p12.1 deletion carriers and age and sex matched controls in the UK Biobank and SPARK were assessed using two-tailed t-tests. The relationship of secondary variant burden and phenotypes in all three cohorts, and comparison with adults and children in the DD cohort, were assessed using two-tailed t-tests. We note that phenotypes in these cohorts were only assessed if they were present in at least five individuals or 10% of the cohort, whichever was larger, and the cohort had at least five or 10% of the cohort controls. We further assessed the effects of secondary variant on phenotypes across cohorts using logistic regression using the model structures *(b)* and *(c)* described above for the DD cohort, without the inclusion of STR variants. Phenotypic logistic regression was performed on main and secondary ICD10 codes with age and sex included as covariates. PheWAS was performed using the *PheWAS* v.0.99.6-1 package in R^114^ on all available samples from the UK Biobank using Phecodes derived from ICD9 and ICD10 codes, identified from Data Fields 41202, 41204, 41203, and 41205 (main and secondary inpatient ICD9/10 codes), while correcting for sex, age, and the top four genetic principal components. Sample sizes, test statistics, uncorrected and corrected p-values, confidence intervals, and variance statistics for all analyses are available in **Table S4**.

### Genotype-phenotype analysis in other neurodevelopmental disease cohorts

We assessed genomic and phenotypic data from individuals from the Simons Searchlight project^63^, ascertained for probands with 16p11.2 deletions and duplications, and Simons Simplex Collection (SSC), ascertained for families with simplex cases of autism^64^. Within the Simons Searchlight cohort, we assessed 128 probands with the 16p11.2 duplication (n=37) or deletion (n=91). Within the SSC cohort, we assessed genomic data of 2,435 total probands, and classified probands with the following primary variant classes for downstream analysis: (i) 1,237 probands with rare, deleterious variants (<0.1% frequency, loss-of-function or missense variants with CADD Phred-like scores >25); (ii) 79 probands with large rare deletions (<0.1% population frequency, >500kbp); (iii) 148 probands with large rare duplications (<0.1% population frequency, >500kbp); and (iv) 1,084 probands who did not carry any of these variants or any other known pathogenic CNVs^33^. We note that groups with primary variants have overlapping samples. Additionally, we assessed phenotypic data for an additional 419 SSC probands and 32 Simons Searchlight probands to compare SRS distributions with 16p12.1 deletion probands (a total of 2,844 total SSC probands and 139 Simons Searchlight probands were used in these analyses). De-identified data from these cohorts were obtained and analyzed according to a protocol approved by the Pennsylvania State University Institutional Review Board (IRB #STUDY00011008). In sum, we assessed secondary variants and phenotypes in 2,252 individuals with primary variants from six cohorts: DD (n=269), SPARK (n=94), MyCode (n=160), UKB (n=250), Searchlight (n=128), and SSC (n=1,351). We also assessed data from 1,084 SSC probands without primary variants. Data from an additional 311,980 control individuals was also included, including non-carrier samples from DD (n=183), age and sex-matched controls without CNVs from SPARK (n=356), PheWAS and age and sex-matched controls without CNVs from UKB (n=310,990), 16p11.2 CNV samples from Searchlight and SSC probands without genetic data but with SRS data (n=32 and 419, respectively).

Exome sequencing VCFs and raw microarray data for Searchlight cohorts, whole genome sequencing VCFs and STR calls for SSC^75^, and all phenotype data were downloaded through the SFARI Base portal (https://www.base.sfari.org). We processed and filtered exome and WGS-based SNVs and indels for the same quality control filters used to process the DD cohort, and then annotated variants using our standardized pipeline. Short tandem repeat calls from SSC were previously published by Mitra and colleagues^75^ and were processed and filtered with the same pipeline as our cohort. CNV calls from microarrays for SSC were previously published by Sanders and colleagues^115^, while CNV calls from microarrays for the Searchlight cohort were processed using PennCNV^89^ as previously described^28^. For this manuscript, genes within CNVs were reannotated using GENCODE v19^85^, but otherwise used as-is without additional processing. Primary variant SNVs and CNVs were removed from secondary variant lists for downstream processing. We further processed microarray data and calculated PRS for both cohorts using the same pipelines as the DD cohort, except that autism PRS was not calculated in SSC samples, as the underlying GWAS summary statistics were calculated in part using SSC samples^41^. Finally, we curated results of quantitative phenotypic assessments for each cohort from SFARI Base, including full-scale IQ, internalizing and externalizing behavior (ABCL/CBCL), social responsiveness (SRS), autism-related behaviors (BSI, Searchlight only), repetitive behavior (RBS-R, SSC only), coordination disorder (DCDQ, SSC only), BMI z-score, and head circumference z-score (Searchlight only).

Linear regression models for assessing variation in these phenotypes were constructed using the *OLS* function from statsmodels v0.14.2, using the same model structures *(b)* and *(c)* described above for the DD cohort (note that Searchlight models did not include STRs). For models investigating the interactions of primary and secondary variants, Benjamini-Hochberg FDR correction was performed using the statsmodels v.0.14.2 *false_discovery_control* function. All other correlations, statistical analyses, GO enrichments, and multiple testing corrections for comparing variant classes and quantitative phenotypes were performed in the same manner as for the DD cohort. Sample sizes, test statistics, uncorrected and corrected p-values, confidence intervals, and variance statistics for all analyses are available in **Table S5**.

### Data and code availability

Whole genome sequencing and SNP microarray data generated in this study are available at NCBI dbGaP phs002450.v2.p1. All code generated for this project, including pipelines for running bioinformatic software and custom analysis scripts, are available at https://github.com/girirajanlab/16p12_WGS_project. Statistical analyses and experimental results for the data presented in **Figs. 2-7** and associated supplementary figures are available in **Tables S2-S6.**

## SUPPLEMENTAL FIGURES

**Figure S1.**
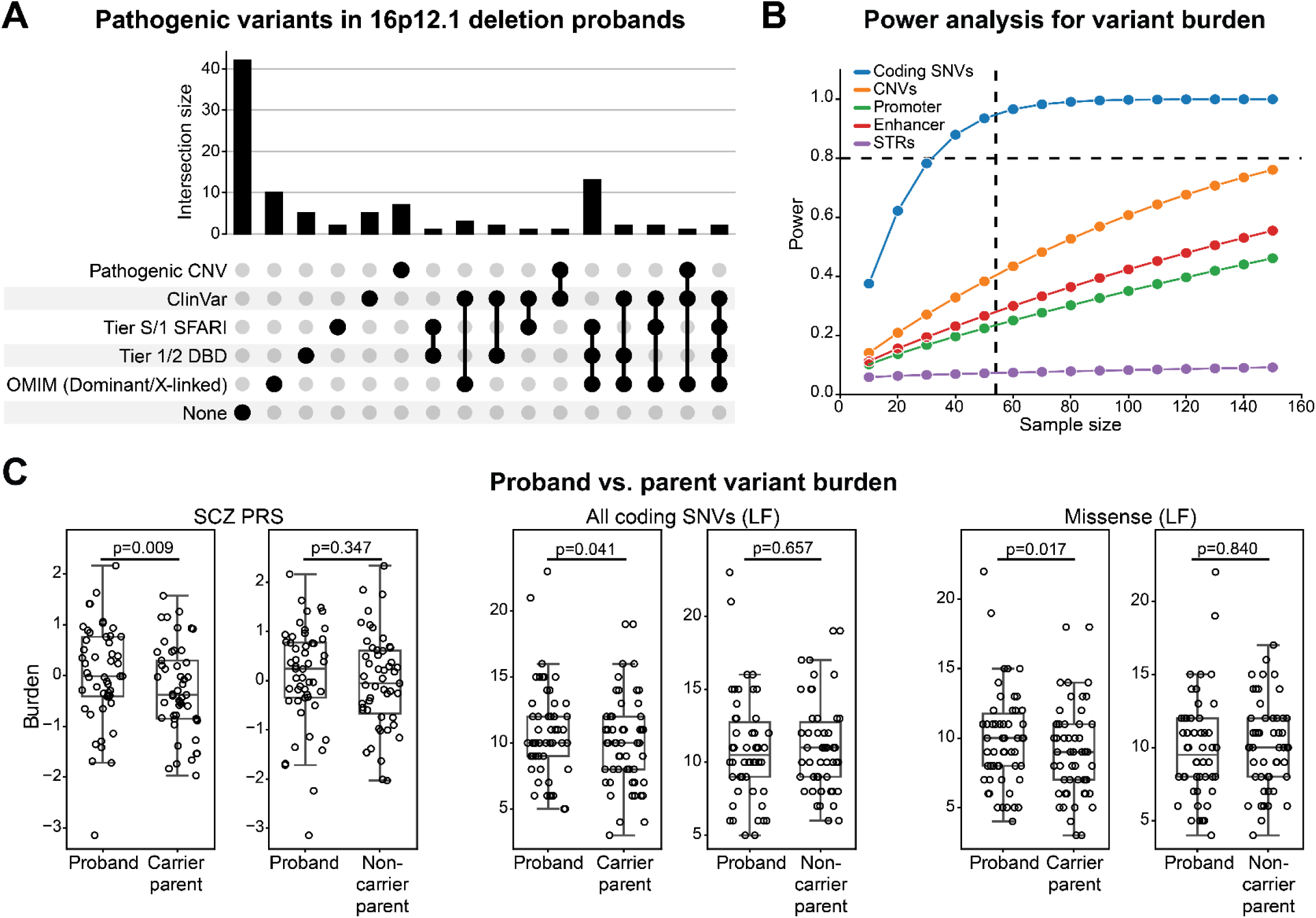
Secondary variant burden comparisons among 16p12.1 deletion family members (related to Figure 3). (**A**) UpSet plot shows the number of 16p12.1 deletion probands with secondary variants in one or more disease-associated categories, potentially indicative of multiple genetic diagnoses. **(B)** Power analysis for detecting changes in burden of rare variant classes among individuals with the 16p12.1 deletion (see Methods). Dashed horizontal line indicates 80% power and dashed vertical line represents the sample size of proband-carrier parent pairs (n=54). **(C)** Changes in burden of SCZ PRS (left), missense (LF) variants (center), and all coding SNVs (LF) (right) between 16p12.1 deletion probands and their carrier (left, n=49-54) and noncarrier parents (right, n=50-51). P-values from one-tailed (rare variants) or two-tailed (SCZ PRS) t-tests.

**Figure S2.**
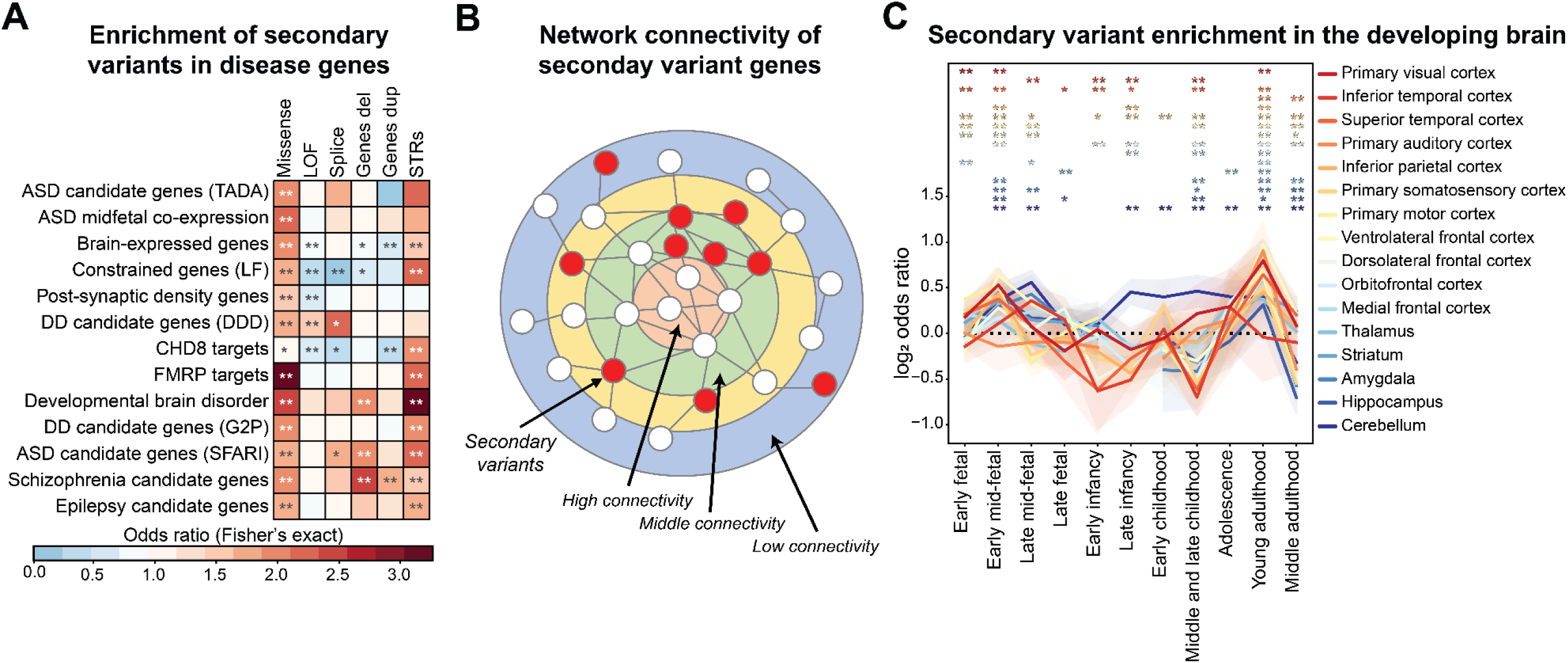
Functional effects of secondary variants observed in 16p12.1 deletion probands (related to Figure 3). (**A**) Enrichment of secondary variant classes in 16p12.1 deletion probands for sets of genes involved with neurodevelopmental disease and related functions. Fisher’s exact test, *p≤0.05, **Benjamini-Hochberg FDR≤0.05. **(B)** Diagram illustrating the distribution of secondary variants (red nodes) in genes with varying connectivity (colored rings) in a brain-specific interaction network. Highly connected genes (light red ring) are depleted for secondary variants, while genes with intermediate connectivity (light green ring) are enriched for variants. **(C)** Line plot shows enrichment (log-odds ratios with 95% confidence intervals; y-axis) of secondary variants in 16p12.1 deletion probands among genes preferentially expressed in 16 brain tissues (colored lines) over 11 developmental timepoints (x-axis). Fisher’s exact test, *p≤0.05, **Benjamini-Hochberg FDR≤0.05.

**Figure S3.**
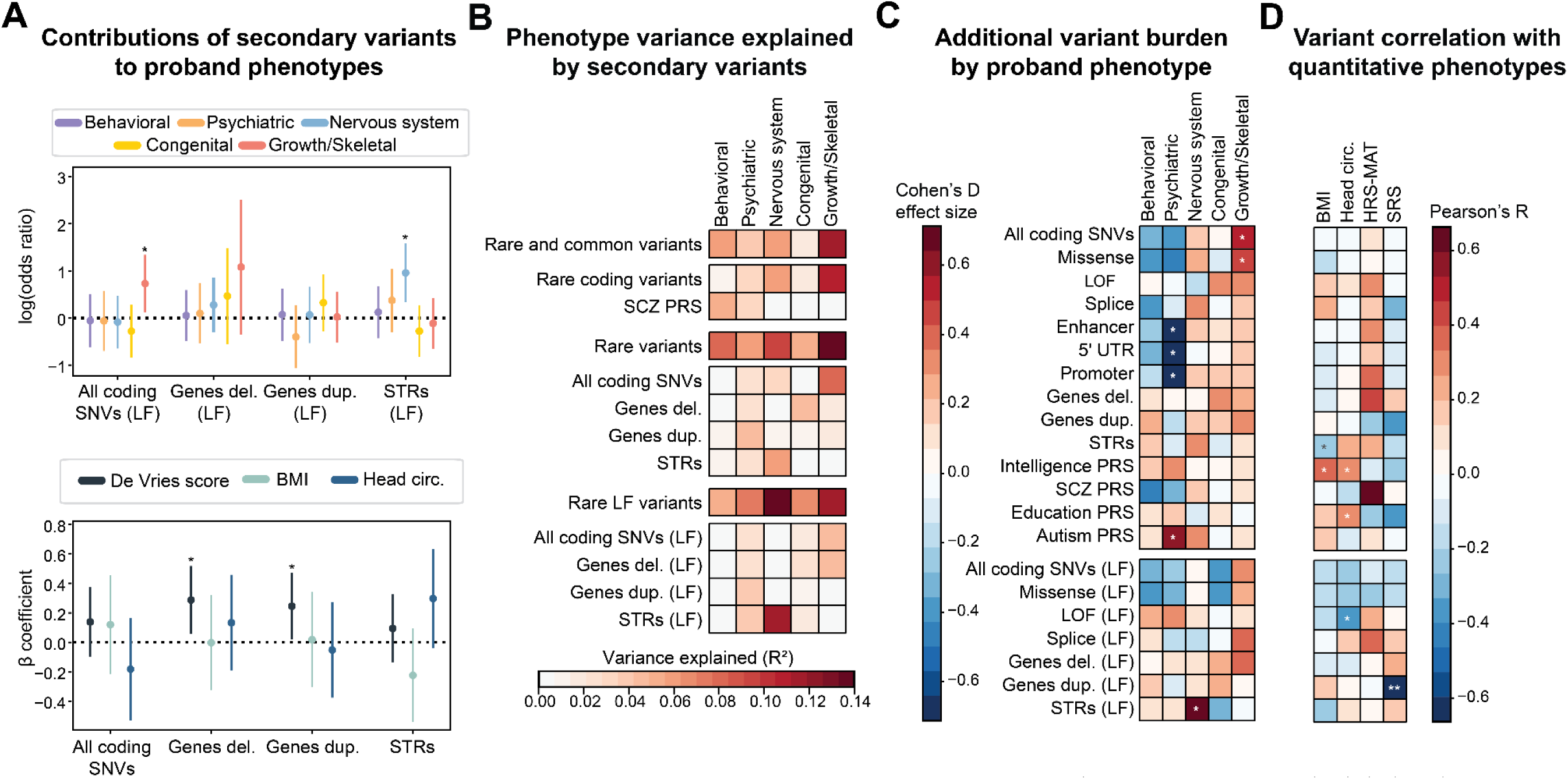
Secondary variant associations with 16p12.1 deletion phenotypic domains (related to Figure 4). (**A**) (Top) Forest plots show log-scaled odds ratios from logistic regression models for secondary variant burden in constrained genes for probands (n=47-71) with higher complexity scores in five phenotypic domains, compared with probands with lower complexity scores for each domain. *p≤0.05. (Bottom) β coefficients from linear regression models for quantitative phenotypes in probands (n=43-76). *p≤0.05. **(B)** Variance explained by secondary variant burden from logistic regression models (McFadden’s pseudo-R^2^), both for individual variant classes and joint contributions from combinations of classes. **(C)** Comparisons of secondary variant burden between 16p12.1 deletion probands (n=53-84) with higher and lower complexity scores for each phenotypic domain. Two-tailed t-test, *p≤0.05, **Benjamini-Hochberg FDR≤0.05. **(D)** Pearson correlations between quantitative phenotypes and secondary variant burden in deletion probands (n=9-59). *p≤0.05, **Benjamini-Hochberg FDR≤0.05.

**Figure S4.**
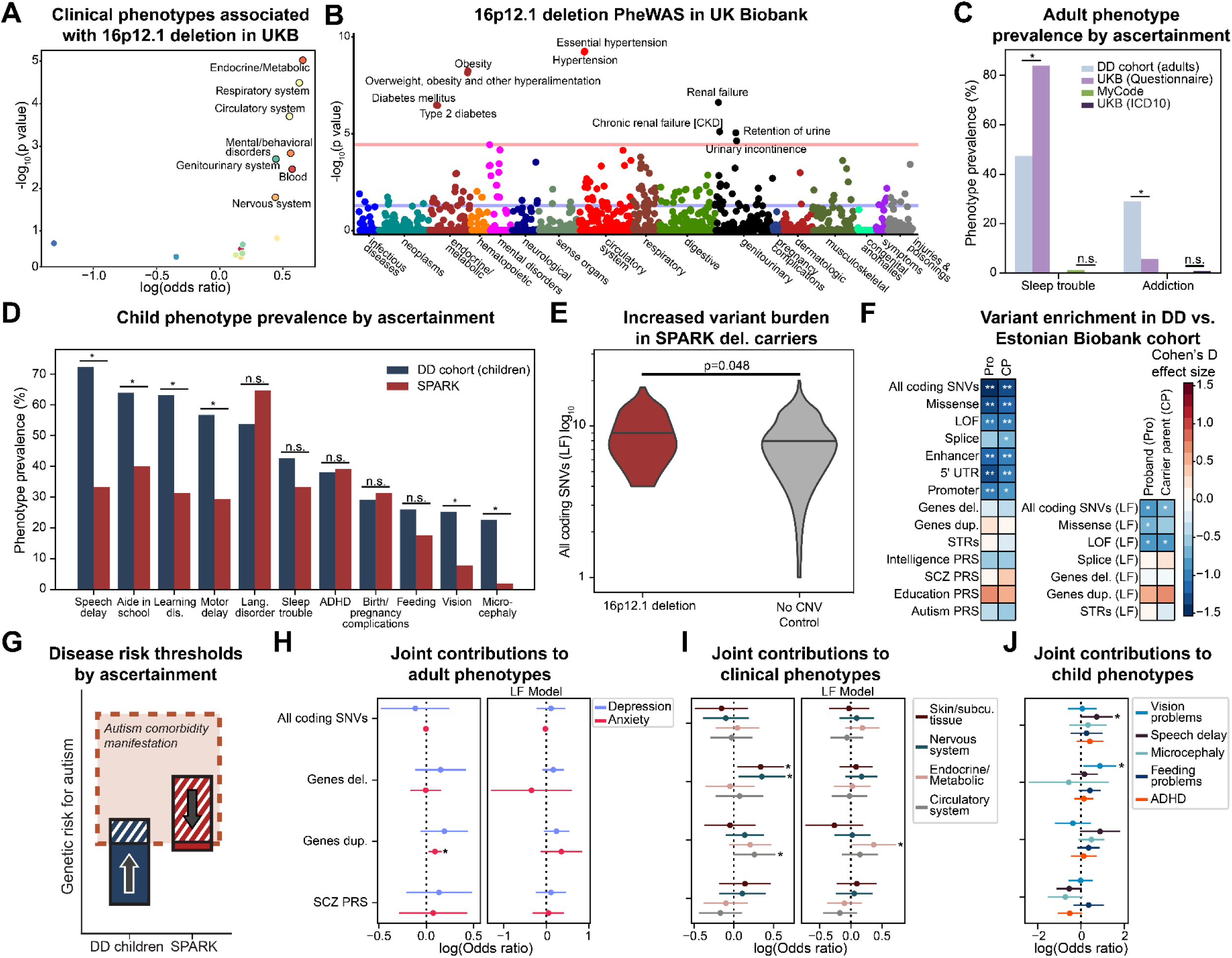
Effects of ascertainment on phenotypes and secondary variant associations in = 16p12.1 deletion carriers (related to Figure 5). (**A**) Enrichment of select ICD10 chapters from logistic regression models in UK Biobank (UKB) 16p12.1 deletion carriers compared to controls without large rare CNVs (n=3,488). P-values from logistic regression. Labeled points indicate phenotypes with Benjamini-Hochberg FDR≤0.05. **(B)** PheWAS analysis for 16p12.1 deletion carriers in UKB (n=99,363-255,262). Colored circles indicate individual phenotype membership in respective ICD10 chapters. Red line indicates phenome-wide significance (Bonferroni p=0.05) and blue line indicates nominal significance (p=0.05). **(C)** Comparison of sleep disturbance and addiction phenotype prevalence in adults from the DD cohort (n=38) and individuals from UK Biobank (questionnaire n=35-249, ICD10 n=217) and MyCode (n=160). *p≤0.05, Fisher’s Exact test. **(D)** Prevalence of developmental and psychiatric phenotypes in children with 16p12.1 deletion from the DD (n=80-151) and SPARK (n=40-51) cohorts. Phenotypes shown were restricted to those present in >20% of probands in either cohort. *p≤0.05, Fisher’s Exact test. **(E)** Comparison of SNV (LF) burden in SPARK individuals with 16p12.1 deletion (n=89, left) to age and sex-matched controls without large rare (>500kb) CNVs (n=356, right). P-value from two-tailed t-test. **(F)** Changes in secondary variant burden between probands (“Pro”, n=97-99) and carrier parents (“CP”, n=54-57) in the DD cohort with 16p12.1 deletion individuals the Estonian Biobank (n=5-8). Blue indicates a depletion in secondary variant burden for Estonian Biobank deletion carriers. One-tailed t-test, *p≤0.05, **Benjamini-Hochberg FDR≤0.05. **(G)** Schematic outlining the proposed relationship between different genetic risk factors in individuals with 16p12.1 deletion across different ascertainments. In cohorts where a majority of participants have a particular disorder, such as autism in SPARK, established risk factors (such as autism PRS) may not show the expected correlations for comorbid features. However, these correlations would be observed in cohorts with different ascertainments (such as the DD cohort). **(H-J)** Forest plots show associations of secondary variants in all genes and constrained genes (“LF Model”) with select phenotypes from joint logistic models in **(H)** DD cohort adults, UK Biobank, and MyCode individuals for psychiatric features (n=331); (**I**) UK Biobank and MyCode individuals for clinical phenotypes from EHR data (n=321); and **(J)** children from the DD cohort and SPARK (n=98-125). *p≤0.05. Full results are available in **Table S4G**.

**Figure S5.**
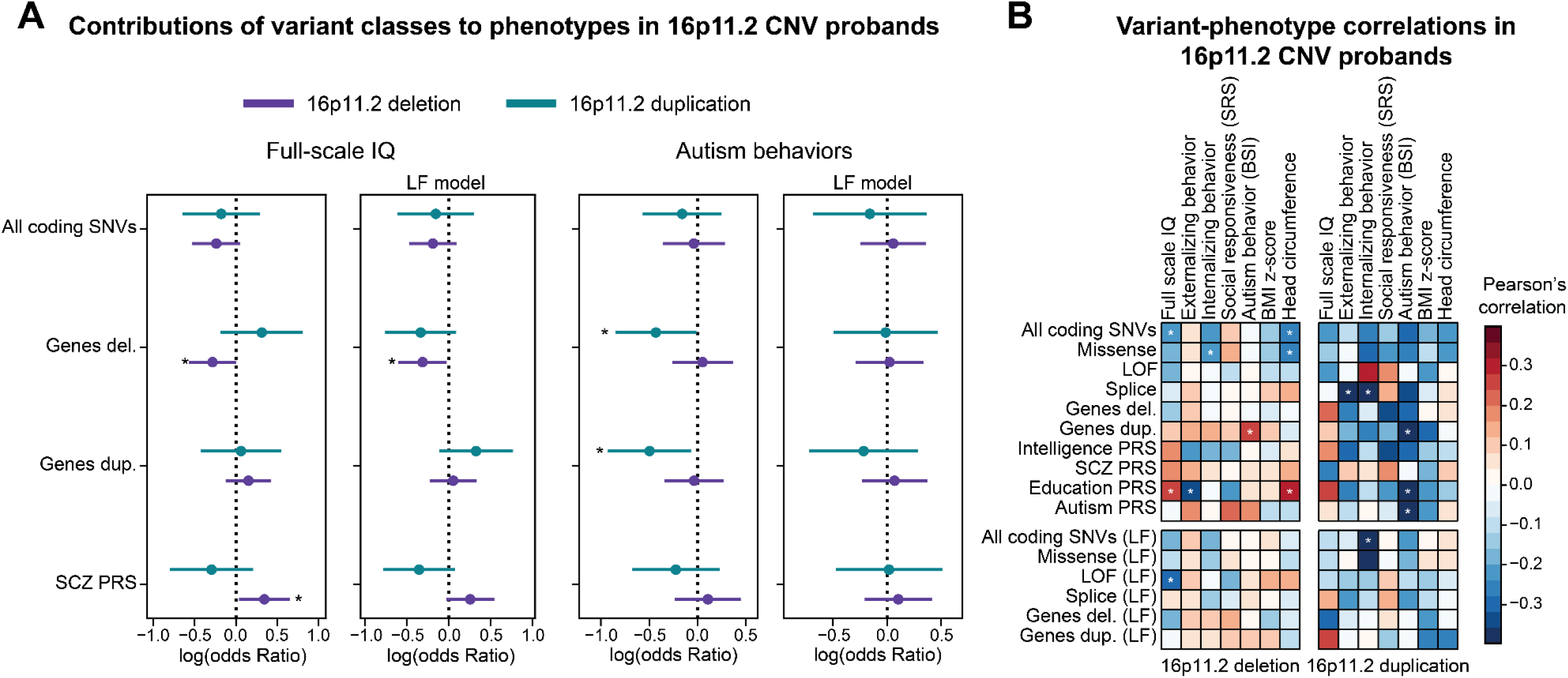
Associations between secondary variants and developmental features of 16p11.2 CNV probands (related to Figure 6). (**A**) Example forest plots show results from select linear regression models for associations between secondary variant classes and full-scale IQ (left) and autism-related behavior (BSI, right) for probands with the 16p11.2 deletion (n=52-57, purple) and duplication (n=21-25, teal). “LF model” indicates models where rare variants are selected for genes under evolutionary constraint. *p≤0.05. Full results are available in **Table S5A**. **(B)** Pearson’s correlations between secondary variant burden and quantitative phenotypes of probands with 16p11.2 deletion (n=58-89) and duplication (n=25-37). *p≤0.05.

**Figure S6.**
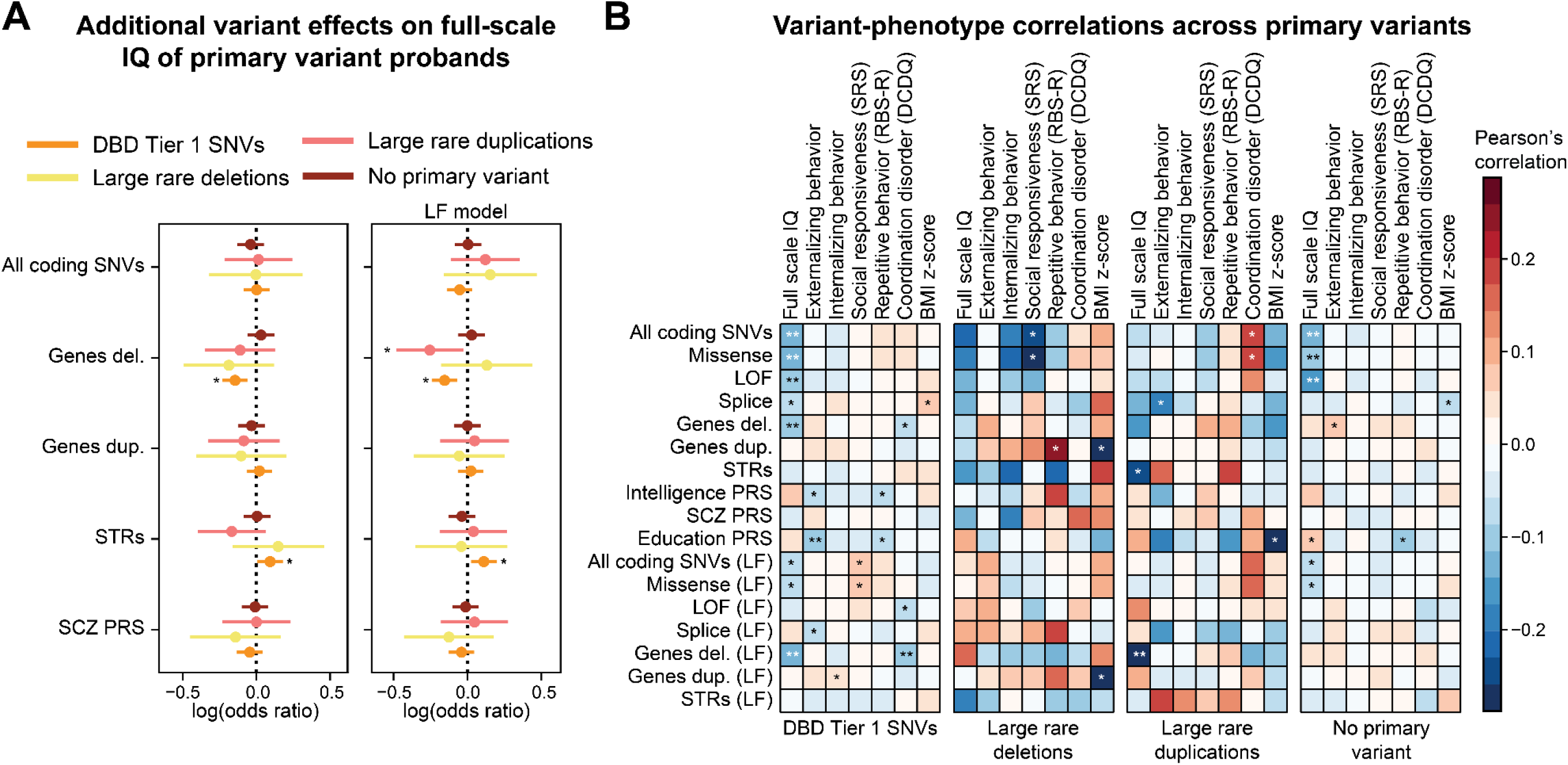
Associations between secondary variants and developmental features of probands with primary variants (related to Figure 6). (**A**) Example forest plots show results from select linear regression models for associations between secondary variant classes and full-scale IQ for probands with pathogenic SNVs in candidate neurodevelopmental genes (n=660, orange), large, rare deletions (n=51, yellow) and duplications (n=85, pink), and probands without such variants (n=632, red) from the SSC cohort. *p≤0.05. “LF model” indicates models where rare variants are selected for genes under evolutionary constraint. Full results are available in **Table S5A**. **(B)** Pearson’s correlations between secondary variant burden and quantitative developmental phenotypes of SSC probands with pathogenic SNVs (n=736-1,236), rare deletions (n=49-78), rare duplications (n=102-148), and probands without such variants (n=671-1,083). *p≤0.05, **Benjamini-Hochberg FDR≤0.05.

## SUPPLEMENTAL TABLES

**Table S1. Description of the DD cohort and phenotypic data (Excel file; related to Figure 1)**. **Table S1A** lists all 452 individuals in the DD cohort, including family relationships, age, biological sex, and 16p12.1 deletion status if known. The table also lists secondary variant burden (rare variant counts and PRS), complexity scores for phenotypic domains (child and adult), quantitative measures (BMI, head circumference, IQ, and SRS), and age at developmental milestone achievement for all individuals with available data. **Table S1B** contains the scoring rubric used to calculate complexity scores for phenotypic domains in children. **Table S1C** lists minimum age thresholds used for identifying psychiatric features in pediatric family members. **Table S1D** lists pathogenic variants or deleterious variants in genes associated with disease that were identified in 16p12.1 deletion probands.

**Tables S2-S5. Statistical analyses (Excel files)**. All statistics supplementary tables are linked to the analyses presented in specific figures, which are detailed in the first sheet of each file. The tables list sample sizes, statistic test used, effect sizes/odds ratios, confidence intervals, and p-values with and without multiple testing correction, depending on the analysis. Gene set enrichments and gene lists for specific analyses are listed under separate table headings. Additional data (i.e., GO enrichments) are also provided in some of the files, which are described below.

**Table S2. Statistics analysis for Figures 2, 3, S1, and S2 (Excel file)**.

**Table S3. Statistics analysis for Figures 4 and S3 (Excel file). Table S3B** lists enriched GO terms for genes with secondary variants among 16p12.1 deletion probands with the five phenotypic domains.

**Table S4. Statistics analysis for Figures 5 and S4 (Excel file). Table S4A** details how psychiatric phenotypes were matched across questionnaire and EHR (ICD10) datasets across cohorts with different ascertainments.

**Table S5. Statistics analysis for Figures 6, S5, and S6 (Excel file). Tables S6C** and **S6D** list enriched GO terms for genes with secondary variants among SSC probands with primary SNVs and CNVs and without primary variants (S6C), and Searchlight probands with 16p11.2 deletions and duplications (S6D).

## Notes

### Competing Interest Statement

The authors have declared no competing interest.

### Funding Statement

This study was funded by NIH R01-GM121907 and resources from the Huck Institutes of the Life Sciences to S.G. M.J. and C.S. were supported by NIH T32-GM102057. A.T. was supported by NIH T32-LM012415. L.P. was supported by Fulbright Commission Uruguay-ANII. A.R. was supported by grants from the Swiss National Science Foundation 31003A_182632. S.B. was supported by the NIHR Manchester Biomedical Research Centre (NIHR203308).

### Author Declarations

The Institutional Review Boards of Pennsylvania State University and Geisinger Health Systems gave ethical approval for this work.

